# Isolated fetal neural tube defects associate with increased risk of placental pathology: evidence from the Collaborative Perinatal Project

**DOI:** 10.1101/2021.03.16.21253704

**Authors:** Marina White, David Grynspan, Tim Van Mieghem, Kristin L Connor

## Abstract

**Objective:** To compare placental pathology and fetal growth in pregnancies with an isolated fetal neural tube defect (NTD; cases) to those without congenital anomalies (controls). We hypothesised that cases would be at an increased risk of placental pathology and poorer anthropometric outcomes at birth compared to controls

**Methods:** We performed a matched case-cohort study using data from the Collaborative Perinatal Project. Cases (n=74) and controls (n=148) were matched (1:2 ratio) for maternal pre-pregnancy BMI, maternal race, infant sex, gestational age at birth and study site. Primary outcomes were placental characteristics (weight and size measurements, pathology). Secondary outcomes were infant birth outcomes. Subgroup analysis was done by type of NTD (spina bifida, anencephaly or encephalocele), infant sex, and preterm/term delivery. Data were analysed using adjusted generalized linear and nominal logistic regression models. Results are presented as adjusted β or adjusted odds ratio (aOR; 95% confidence interval).

**Results:** Cases had lower placental weight (β=-22.2 g [-37.8 – −6.6]), surface area (β=-9.6 cm^2^ [-18.3 – −1.0]) and birth length z-scores (β=-0.4 [-0.7 – −0.001]) compared to controls. Cases were more likely to have a single umbilical artery (vs. two; 6 [8.1%] vs. 1 [0.7%]; aOR=301 [52.6 – 1726]), overall placental hypermaturity (9 [12.2%] vs. 5 [3.4%]; aOR=6.8 [3.1 – 14.7]), and many (vs. few) Hofbauer cells (9 [12.2%] vs. 7 [4.7%]; aOR=3.02 [1.2 – 7.3]), stromal fibrosis (9 [12.2%] vs. 10 [6.8%]; aOR=3.0 [1.4 – 6.3]) and pathological edema (11 [14.9%] vs. 12 [8.1%]; aOR=3.04 [1.4 – 6.7]) in placental terminal villi compared to controls. Placental pathology varied across NTD subtypes, infant sex, and preterms vs. term pregnancies.

**Conclusions:** Fetuses with isolated NTDs may be at increased risk of placental pathology, which could be contributing to poor fetal growth in these pregnancies and subsequent postnatal morbidities.

## Introduction

Neural tube defects (NTDs) are amongst the most common congenital anomalies and affect more than 300,000 infants annually worldwide^1^. Most NTDs are multifactorial in origin^2^, but folic acid and other micronutrients play an important role in their pathogenesis^3^. Introduction of periconceptional folic acid supplementation and food fortification with folic acid has significantly decreased the incidence of NTDs in countries adhering to these strategies^4^. Despite these initiatives, certain subgroups remain at high risk of carrying a pregnancy with an isolated NTD, including the growing population of people with diabetes^5^ or obesity^6^.

The neurologic severity of NTDs varies depending on their location along the neural tube as well as the degree of central nervous system involvement. Where anencephaly is invariably lethal, spina bifida and (smaller) encephaloceles are compatible with life, but are associated with postnatal morbidity^7^. Beside the impact of NTDs on the infant’s central nervous system function, they are also associated with an increased risk of low birthweight^8,9^, lower gestational age at birth^8,9^ and fetal growth restriction^10^, which may increase the risk of fetal or postnatal mortality.

Placental and umbilical cord maldevelopment, which is a major driver of preterm birth^11^ and fetal growth restriction^12^, has been insufficiently studied in pregnancies with fetal NTDs and no large-scale studies have compared placental development in fetuses with NTDs with healthy controls. Better insight into the development and function of placentae from fetuses with NTDs is nevertheless required, as increased understanding of the mechanisms that drive poor growth and early birth in these fetuses will aid in developing targeted, early interventions to improve their developmental trajectories.

The aim of our study was therefore to assess whether placental pathology is more prevalent in fetuses with isolated NTDs compared to fetuses without any congenital anomalies. We hypothesised that pregnancies carrying a fetus with an isolated NTD would be at an increased risk of placental pathology and poorer anthropometric outcomes at birth compared to control fetuses. We also explored differences in placental and infant birth outcomes related to NTD subtype, infant sex, and between infants born preterm and full term.

## Methods

### Data source, collection and study design

The Collaborative Perinatal Project (CPP) was a prospective cohort study aimed at understanding how biomedical, environmental (socioeconomic factors), and genetic factors interact to influence pregnancy outcomes and child health^13^. The CPP dataset contains longitudinal data (from enrolment during pregnancy to seven years postpartum) from approximately 58,000 pregnancies between 1959 and 1965, from 12 clinical sites in the United States and is publicly available online^13^. Data on maternal characteristics, obstetric and medical history, and socioeconomic demographics were collected at the first prenatal visit. Prenatal observations were recorded at each subsequent visit, and data on labour and delivery events and neonatal outcomes were collected at birth. Placental pathology (gross and microscopic) was assessed using samples collected at the time of delivery. Subsequent detailed clinical assessments of the children were performed at one and at seven years of age.

We used a case-cohort study design to compare prevalence of placental pathology in fetuses with isolated NTDs to a subcohort of healthy control fetuses (with no congenital anomalies), matched for specific maternal/pregnancy characteristics (see below, Figure 1). Data were accessed in April 2020 and data cleaning and case-control matching were done in RStudio (version 3.6.1, PBC, Boston, MA). Select variables of interest were identified from the master file, variable file, and work file 5 (congenital malformations, one and seven years)^13^ and linked using the participant identification number.

**Figure 1.**
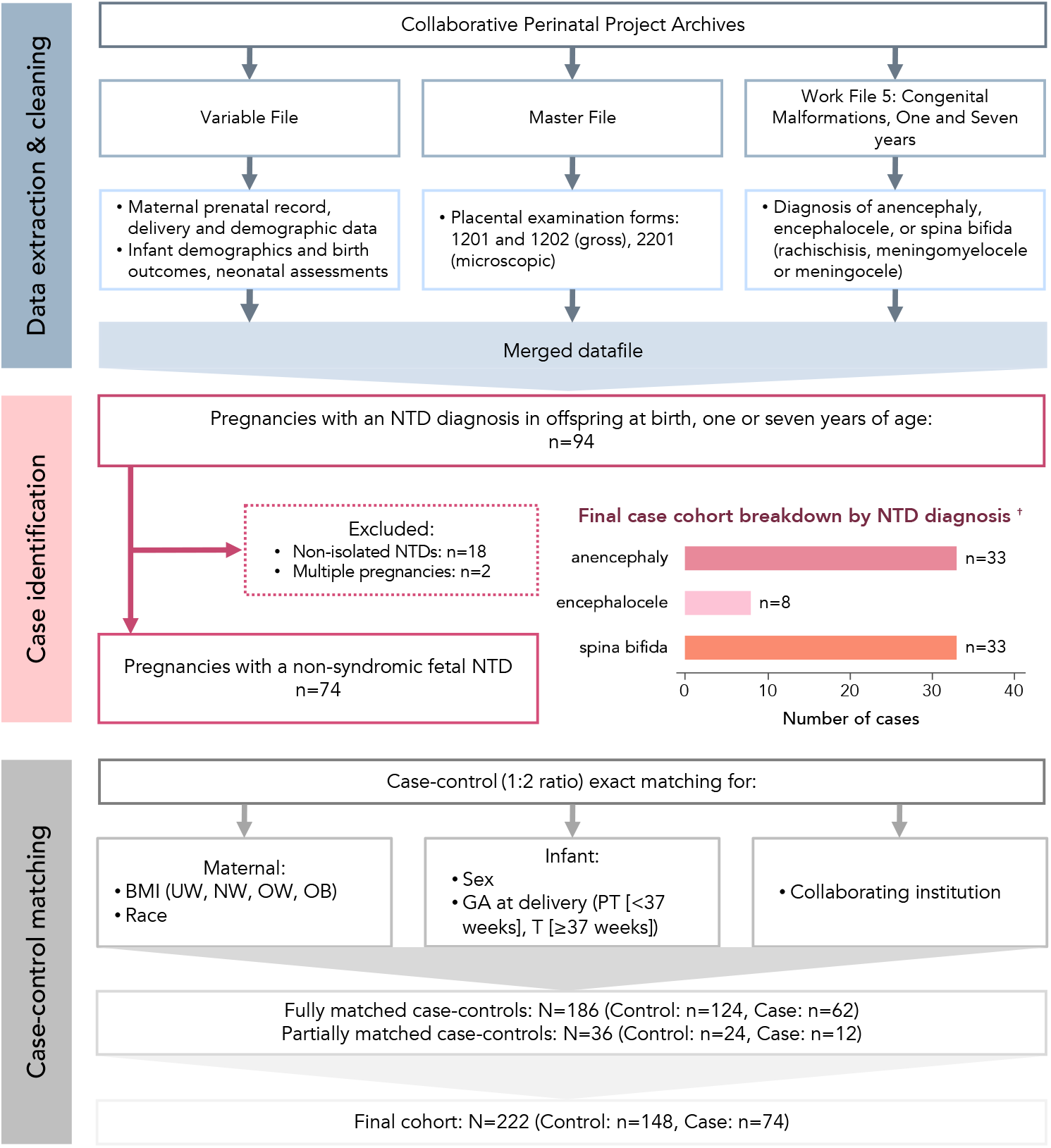
Study flow through data extraction, case identification and classification, and case-control matching. † Where cases were diagnosed with more than one NTD (spina bifida and anencephaly [n=3], spina bifida and encephalocele [n=1]), they have been classified according to the more severe diagnosis (least to most severe: spina bifida, encephalocele, anencephaly). NTD, neural tube defect.

### Case identification

Cases were defined as singleton infants diagnosed with an isolated NTD (anencephaly, encephalocele, spina bifida) at birth, one or seven years of age. Infants with additional congenital disorders indicative of a non-isolated NTD were excluded, as were multifetal pregnancies due to their known higher risk of complications, including placental pathologies^14^.

### Case-control matching

Controls were singleton pregnancies, with available placental pathology data, in whom no congenital anomalies were diagnosed. Controls were matched to cases using MatchIt^15^, an R software package that generates case-control matches using exact covariate matching, in a 1:2 (case:control) ratio for maternal and infant variables with known influence on pregnancy outcomes, including maternal pre-pregnancy BMI (categorized as underweight [<18.5 kg/m^2^], normal weight [18.5-24.9 kg/m^2^], overweight [25.0-29.9 kg/m^2^], obese [≥30.0 kg/m^2^]), maternal race (categorized as Black, White, Puerto Rican, Other), infant sex, gestational age at delivery (categorized as <37 weeks [preterm], ≥37 weeks [full term]) and contributing hospital site.

Maternal race was considered in matching criteria due to the marked disparities in maternal and fetal health outcomes for Black (compared to White) mothers in the United States that are consequences of institutionalized and interpersonal racism^16^ and increased allostatic load^17^. Additionally, high maternal pre-pregnancy BMI was also included in the matching criteria as it is known to effect placental development and function^18^ and increase risk of fetal NTDs^6^. The groups were also matched for infant sex to account for sex-based differences in placental development and function^19^, and because female fetuses are more susceptible to developing an isolated NTD^20^. Lastly, the groups were matched for hospital site from which the mothers were recruited, as each contributing hospital site had its own specific participant sampling frame^13^ and health care access and health status varies widely across the US^21^.

### Infant birth outcomes

Gestational age at birth was estimated using maternal reports of the date their last-menstrual period began. For all analyses, a gestational age >43 weeks at birth was deemed likely implausible and was marked as missing^22^. Infant Apgar scores were collected at one and five minutes postpartum. Anthropometric variables (birth weight, length, head circumference) were transformed to z-scores adjusting for sex- and gestational age using the INTERGROWTH-21^st^ data^23^. For analyses of infant birth anthropometry and Apgar scores, cases with anencephaly were only considered in analyses of birth weight as the lack of cranium and brain will influence infant length. Moreover, as anencephaly is a lethal condition, Apgar scores are likely irrelevant.

### Placental pathology

Data on placental anthropometry (weight, largest and smallest diameter and thickness) and pathology (gross and microscopic) were recorded from examinations of fresh placentae by pathologists blinded to the clinical outcome of the pregnancy following delivery. In addition to raw measures of placental weight and dimensions, we calculated gestational-age corrected placental weight z-scores^24^. Placental surface area (assuming an elliptical surface) was estimated as:^25^

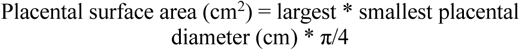

We calculated ratios for infant birth weight-to-placental weight, largest and smallest diameter, and surface area.

The following aspects of placental histopathology were assessed, as per the original cohort study protocol:

1. Pathologies of the umbilical cord: cord edema (present vs. not), single umbilical artery (vs. two), neutrophilic infiltration in the umbilical cord (present [slight, marked or moderate] vs. not seen)
2. Pathologies of the membranes and fetal surface: fetal thrombotic vasculopathy (present vs. not seen), membrane cysts (present vs. not seen), neutrophilic infiltration of the amnion or chorion of the membrane roll and placental surface (present [slight, marked or moderate] vs. not seen)
3. Pathologies of the maternal surface: infarcts (present vs. none), calcifications (none, maternal surface only, throughout)
4. Villous pathology: presence of Langhans’ layer (cytotrophoblast cell layer; present vs. not seen), pathological edema (present vs. not seen), stromal fibrosis (present vs. not seen), and Hofbauer cells (fetal tissue macrophages; few vs. many). Syncytial knots, which are also an indicator of placental maturity and increase across gestation^26^, were only assessed in term (≥37 weeks’ gestation) placentae.

Overall assessment of placental maturity (<20, 20-27, 28-36, or ≥37 weeks’ gestation). We compared this estimate against the gestational age at delivery of each infant to classify their placenta as: immature, appropriate maturity, or hypermature.

### Maternal characteristics

Maternal demographic (age, race, education level, annual family income, marital status, smoking history) and pregnancy (self-reported pre-pregnancy weight, measured height, gestational weight change, diabetes mellitus status and gravidity) characteristics were obtained. Maternal diabetes status was classified as not present, or present before pregnancy and/or during pregnancy and/or postpartum. Due to the rarity of any diabetes mellitus during the perinatal period (4.5% of our cohort), and the lack of available data on type of diabetes mellitus, we collapsed these data into any vs. no evidence of diabetes during the perinatal period. A numerical socioeconomic indicator (range 0 [lowest] to 9.5 [highest]) was also reported using each household’s chief earner’s education level and occupation and total family income, as described elsewhere27. Data on maternal smoking was provided as number of cigarettes per day, which we categorized as non-smokers, light smokers (<1 pack/day or <20 cigarettes) and heavy smokers (≥1 pack/day or ≥20 cigarettes).

### Statistical analyses

Data were analysed using JMP (version 14.0, SAS Institute Inc, Cary, NC). The Wilcoxon Rank Sum test and one-way Analysis of Variance (ANOVA) were used to test differences in medians (non-parametric data) and means (parametric data), respectively, for maternal cohort characteristics, and infant and placental birth outcomes between case and control groups. Associations between study groups and categorical outcome variable distribution were tested using the Likelihood Ratio Chi Square test. We also performed adjusted analyses of relationships between infant study group and infant and placental birth outcomes using generalized linear models^28^. Maternal age (13-43 years; continuous), smoking (non-, light- or heavy smoker; categorical), socioeconomic index (0-9.5; continuous), diabetes during the perinatal period (present vs. absent), and infant gestational age at delivery (22-43 weeks; continuous) were included in the model. Continuous data are reported as median (IQR) or mean ± SD (β [95% CI]) and p values from ANOVA/Wilcoxon (unadjusted) and generalized linear models (adjusted). Categorical data are n (%; Kendall’s τ [95% CI]) with p values from Likelihood Ratio Chi Square test.

Relationships between isolated fetal NTDs and placental pathology (present vs. not seen) were assessed using unadjusted and adjusted (as described above) nominal logistic regression models. Participants were weighted using the inverse of the sampling fraction^29,30^. These data are reported as unadjusted odds ratio (OR) or adjusted OR (aOR) with 95% CI and p value from Likelihood Ratio Chi Square test.

Analyses of infant birth outcomes and placental characteristics were also stratified according to type of NTD (spina bifida, anencephaly or encephalocele). Further, as placental morphology and pathology vary widely, and are informed by gestational age^31^, we also explored placental outcomes for case vs. control preterm (<37 weeks’ gestation) and term (≥37 weeks’ gestation) infants separately. Lastly, we investigated whether there were sex-based differences in infant and placental anthropometry (Wilcoxon Rank Sum/ANOVA) or placental pathology (Likelihood Ratio Chi Square). Results below are presented as measure of effect (95% CI) with p values from adjusted analyses.

## Results

### Cohort selection and baseline characteristics

We identified 94 children who had a diagnoses of one or more NTDs (Figure 1). Twenty were excluded due to associated anomalies (n=18) or multiple gestation (n=2; Supplementary Table 1), resulting in a total cohort of 74 cases. Of these, there were 33 cases of spina bifida (45%), 33 cases of anencephaly (45%) and 8 encephaloceles (11%).

Exact matches in a 1:2 (case-control) ratio for the complete matching criteria were achievable for 62 cases. The remaining 12 cases were missing data on maternal BMI, and of these, 11 were matched for the remaining four criteria. The single remaining case was matched to two controls for: maternal race, infant sex, and hospital site. The final cohort size was N=222, with 74 cases and 148 controls (Figure 1).

Demographic characteristics of the final cohort are presented in Table 1. No significant differences were seen between cases and controls for maternal demographic or pregnancy characteristics.

**Table 1.** Cohort characteristics of mothers of infants with isolated neural tube defects and matched controls.

| Maternal characteristics | Control (n=148) | Case (n=74) | p value |
| --- | --- | --- | --- |
| Age (years) | 23 (20 – 30) | 23.5 (20 – 28) | 0.73 |
| Race (n [%]) |  |  | 1 |
| Black | 46 (31.1) | 23 (31.1) |  |
| White | 84 (56.8) | 42 (56.8) |  |
| Puerto Rican | 16 (10.8) | 8 (10.8) |  |
| Other | 2 (1.4) | 1 (1.4) |  |
| Education (years) | 11 (9 – 12) | 10 (9 – 12) | 0.81 |
| Education grouped (n [%]) |  |  | 0.77 |
| Less than high school ($\leq 9$ years) | 47 (31.8) | 21 (28.4) | |
| High school (10 – 12 years) | 84 (56.8) | 45 (60.8) |  |
| College and above ($> 12$ years) | 15 (10.1) | 6 (8.1) | |
| Unknown | 2 (1.4) | 2 (2.7) |  |
| Annual family income (n [%])<sup>†</sup> |  |  | 0.35 |
| $\leq \$1,999$ | 14 (9.5) | 12 (16.2) | |
| \$2000–4999 | 77 (52) | 39 (52.7) | |
| \$5000–7999 | 35 (23.7) | 12 (16.2) | |
| $\geq \$8,000$ | 10 (6.8) | 4 (5.4) | |
| Unknown | 12 (8.1) | 7 (9.5) |  |
| Socioeconomic index (n [%]) |  |  | 0.69 |
| 0.0–1.9 | 8 (5.4) | 8 (10.8) |  |
| 2.0–3.9 | 46 (31.0) | 23 (31.1) |  |
| 4.0–5.9 | 46 (31.1) | 20 (27) |  |
| 6.0–7.9 | 29 (19.6) | 14 (18.9) |  |
| 8.0–9.5 | 14 (9.5) | 6 (8.1) |  |
| Unknown | 5 (3.4) | 3 (4.1) |  |
| Marital status |  |  | 0.89 |
| Single | 20 (13.5) | 11 (14.9) |  |
| Married/common law | 118 (79.7) | 57 (77) |  |
| Widowed/divorced/separated | 10 (6.8) | 6 (8.1) |  |
| Smoking history (n [%]) |  |  | 0.79 |
| Non-smoker | 84 (56.8) | 38 (51.4) |  |
| Light smoker ($< 1$ pack/day) | 38 (25.7) | 20 (27) | |
| Heavy smoker ($\geq 1$ pack/day) | 26 (17.6) | 15 (20.3) | |
| Unknown |  | 1 (3.7) |  |
| Pre-pregnancy weight (kg) | 58.1 (50.8 – 63.5) | 59.0 (53.5 – 65.8) | 0.22 |
| Pre-pregnancy BMI classified (n [%]) |  |  | 0.38 |
| Underweight ($< 18.5$ kg/m<sup>2</sup>) | 18 (12.2) | 4 (5.41) | |
| Normal weight (18.5–24.9 kg/m<sup>2</sup>) | 90 (60.8) | 42 (56.8) |  |
| Overweight (25.0–29.9 kg/m<sup>2</sup>) | 26 (17.6) | 13 (17.6) |  |
| Obese ($\geq 30.0$ kg/m<sup>2</sup>) | 13 (8.8) | 3 (4.1) | |
| Unknown | 1 (0.7) | 12 (16.2) |  |
| Gestational weight change |  |  |  |
| Weight gain (kg) | 10 (7.7 – 12.6) | 9.07 (6.8 – 13.2) | 0.30 |
| Weight loss (n [%]) | 2 (1.4) | 0 | – |
| Unknown | 2 (1.4) | 7 (9.5) |  |
| Diabetes mellitus (n [%]) | 5 (3.8) | 5 (6.8) | 0.27 |
| Gravidity | 2 (0 – 4) | 2 (0.5 – 4) | 0.87 |

<sup>†</sup> One United States Dollar (USD) in 1959 is equal to approximately 8.90 USD in 2020 (US Bureau of Labor Statistics: CPI Inflation Calculator). The approximate equivalent categories for family income (rounded to the nearest hundred dollars) in 2020 would be: $\leq \$17,800$ , $\$17,801$ to $\$44,500$ , $\$44,501$ to $\$71,200$ , $\geq \$71,201$ .
Continuous data are median (IQR; Wilcoxon test to test equality of medians for non-parametric data) or n (%; Likelihood Ratio Chi Square test to test associations between study groups and categorical variables).

### Pregnancy, delivery and neonatal outcomes

Gestational age at birth was marked as missing for the seven controls and 10 cases with a gestational age that was >43 weeks. Mean gestational age at birth was similar between cases and controls, but the incidence of extreme and very preterm birth was higher in cases (Table 2). Birth before 32 weeks’ gestation occurred in 14.9% of cases and 3.4% of controls (p=0.005).

**Table 2.** Birth outcomes for infants with isolated neural tube defects and matched controls.

| Infant characteristics | Control (n=148) | Case (n=74)<sup>†</sup> | Unadjusted β (95% CI) | Adjusted β (95% CI)<sup>‡</sup> |
| --- | --- | --- | --- | --- |
| Male-to-female sex ratio | 0.7 | 0.7 | - | - |
| GA at birth (weeks)<sup>§</sup> | 39 (36 – 41) | 39 (34 – 40) | -0.8 (-1.4 – 0.1) | -0.5 (-1.1 – 0.1) |
| <37 weeks GA at birth (n [%]) | 46 (31.1) | 24 (32.4) |  | - |
| Sub-categories of preterm birth (n [%]) |  |  | -0.4 (-0.6 – -0.2)**<sup>†</sup> | - |
| Moderate/late preterm (32-36 weeks GA) | 41 (27.7) | 13 (17.6) |  |  |
| Very preterm (28-32 weeks GA) | 1 (0.7) | 3 (4.1) |  |  |
| Extremely preterm (<28 weeks GA) | 4 (2.7) | 8 (10.8) |  |  |
| zWeight at birth<sup>†</sup> | 0.4 (-0.7 – 1.3) | -0.4 (-0.9 – 1.3) | -0.2 (-0.5 – 0.1) | -0.2 (-0.5 – 0.1) |
| zLength at birth | 1 ± 1.5 | 0.2 ± 2.1 | -0.4 (-0.8 – -0.02)* | -0.4 (-0.7 – -0.001)* |
| zHead circumference at birth | 0.4 (-0.4 – 1.3) | 0.1 (-0.7 – 1.7) | 0.2 (-0.2 – 0.5) | 0.1 (-0.3 – 0.5) |
| Apgar at one minute (/10) | 8 (7 – 9) | 5 (2 – 8) | -1.5 (-2.2 – -0.8)*** | -1.4 (-2.2 – -0.6)** |
| Apgar at five minutes (/10) | 9 (9 – 10) | 8 (2 – 9) | -1.3 (-1.6 – -0.9)*** | -1.3 (-1.6 – -0.9)*** |

<sup>†</sup> Cases with anencephaly (n=33) were excluded from analyses of birth length and head circumference and Apgar scores. <sup>‡</sup> Adjusted for maternal age, smoking, socioeconomic status, gestational age at birth, and maternal diabetes status in pregnancy (control: n=136, case: n=54). <sup>§</sup> Gestational age at birth was marked as missing for infants with a gestational age that was >43 weeks (controls: n=7, cases: n=10). <sup>†</sup> Effect estimate is Kendall's τ (95% CI).
Continuous data are median (IQR) or mean ± SD with parameter estimates (β [95% CI]) and *p* values from unadjusted (Wilcoxon test to test equality of medians for non-parametric data, ANOVA to test equality of means for normal data with equal variance) and adjusted (Generalized linear models) analyses. Categorical data are n (%) with Kendall's τ (95% CI) and *p* values from Nominal Logistic regression models (Likelihood Ratio Chi Square test to test associations between study groups and variable distributions). For analyses of infant birth anthropometry (standardized for gestational age- and sex), gestational age at birth or distribution of sub-categories of preterm birth, gestational age at birth was not included. Statistical significance is denoted as \**p*<.05, \*\**p*<.01, \*\*\**p*<.001. GA, gestational age.

Overall, cases also had lower length z-scores at birth (β=-0.4 [-0.7 – − 0.001], p=0.049) and Apgar scores at one (β=-1.4 [-2.2 – −0.6], p=-0.001) and five (β=-1.3 [-1.6 – −0.9], p<.0001) minutes compared to controls (Table 2). There were no differences between cases and controls for other infant anthropometry at birth.

In subgroup analyses comparing case and control birth outcomes stratified by NTD subtype, cases with spina bifida had a higher incidence of extremely preterm birth (<28 weeks; 18.2% vs. 3.03%, p=0.003), lower length z-scores at birth (β=-0.5 [-1.0 – −0.04], p=0.03), and lower Apgar scores at one (β=-0.9 [-1.5 – −0.4], p=0.03) and five (β=-0.5 [-1.01 – −0.1], p=0.03) minutes compared to controls (Table 3). Cases with anencephaly had younger gestational age at birth (β=-0.8 [-1.6 – −0.1], p=0.03) and lower birth weight z-scores (β=-0.9 [-1.5 – −0.3], p=0.003) than controls (Table 3). There were no differences in birth outcomes for cases with encephalocele compared to matched controls (Supplementary Table 2).

**Table 3.** Infant birth outcomes for cases with spina bifida or anencephaly in comparison to matched controls.

| Infant characteristics | Control (n=66) | Spina bifida (n=33) | Unadjusted $\beta$ (95% CI) | Adjusted $\beta$ (95% CI) <sup>†</sup> | Control (n=66) | Anencephaly (n=33) | Unadjusted $\beta$ (95% CI) | Adjusted $\beta$ (95% CI) <sup>†</sup> |
| --- | --- | --- | --- | --- | --- | --- | --- | --- |
| Male-to-female sex ratio | 0.65 | 0.65 | - | - | 0.57 | 0.57 | - | - |
| GA at birth (weeks) <sup>‡</sup> | 39 (36 – 41) | 39.5 (34.8 – 40.3) | -0.6 (-1.7 – 0.4) | -0.4 (-1.4 – 0.6) | 39 (35.8 – 41) | 37 (34 – 40) | -0.9 (-1.7 – -0.08)** | -0.8 (-1.6 – -0.1)* |
| <37 weeks GA at birth (n [%]) | 18 (27.3) | 9 (27.3) |  |  | 24 (36.4) | 13 (39.4) |  |  |
| Sub-categories of PTB (n [%])<sup>§</sup> |  |  | -1.3 (-2.3, -0.4)**<sup>‡</sup> | - |  |  | -0.9 (-3.4, 0.6)<sup>‡</sup> | - |
| Moderate/late preterm | 15 (22.7) | 3 (9.1) |  |  | 24 (36.4) | 9 (27.3) |  |  |
| Very preterm | 1 (1.5) | 0 |  |  | 0 | 3 (9.1) |  |  |
| Extremely preterm | 2 (3.03) | 6 (18.2) |  |  | 0 | 1 (3.03) |  |  |
| zWeight at birth | 0.4 (-0.8 – 1.3) | -0.5 (-0.9 – 1.5) | -0.004 (-0.5 – 0.5) | -0.1 (-0.6 – 0.4) | 0.4 (-0.5 – 1.8) | -0.9 (-2.9 – -0.4) | -1 (-1.6 – -0.4)* | -0.9 (-1.5 – -0.3)** |
| zLength at birth | 0.7 (-0.1 – 1.7) | 0.4 (-1.5 – 1.5) | -0.4 (-0.9 – 0.1) | -0.5 (-1.0 – -0.04)* | - | - | - | - |
| zHead circumference at birth | 0.4 (-0.8 – 1.1) | -0.02 (-0.8 – 1.5) | 0.1 (-0.4 – 0.6) | 0.1 (-0.4 – 0.6) | - | - | - | - |
| Apgar at one minute (/10) | 8 (7 – 9) | 6.5 (4 – 8.8) | -0.8 (-1.3 – -0.2)* | -0.9 (-1.5 – -0.4)** | - | - | - | - |
| Apgar at five minutes (/10) | 9 (9 – 10) | 9 (8 – 9.8) | -0.4 (-0.9 – 0.03)* | -0.5 (-1.01 – -0.1)* | - | - | - | - |

<sup>†</sup> Adjusted for maternal age, smoking, socioeconomic status, gestational age at birth, and maternal diabetes status in pregnancy. For analyses of infant birth anthropometry (standardized for gestational age- and sex), gestational age at birth or distribution of sub-categories of preterm birth, gestational age at birth was not included. <sup>‡</sup> Gestational age at birth was marked as missing for infants with a gestational age that was >43 weeks (cases with spina bifida: n=2, matched-controls: n=2; cases with anencephaly: n=6, matched-controls: n=3). <sup>§</sup> Moderate/late preterm: 32-36 weeks' gestation, very preterm: 28-32 weeks' gestation, extremely preterm: <28 weeks' gestation. <sup>‡</sup> Effect estimate is Kendall's $\tau$ (95% CI).
Continuous data are median (IQR) with parameter estimates ( $\beta$ [95% CI]) and *p* values from unadjusted (Wilcoxon test to test equality of medians) and adjusted (Generalized linear models) analyses. Categorical data are n (%) with Kendall's $\tau$ (95% CI) and *p* values from Nominal Logistic regression models (Likelihood Ratio Chi Square test to test associations between study groups and variable distributions). Statistical significance is denoted as \**p*<.05, \*\**p*<.01, \*\*\**p*<.001. GA, gestational age. PTB, preterm birth.

### Placental anthropometry and pathology

There were 7 and 9 cases with missing gross or microscopic placental examinations, respectively. Cases had lower measured placental weight (β=-22.2 [-37.8 – −6.6], p=0.006) and gestational-age standardized placental weight (β=-0.4 [-0.8 – −0.04], p=0.009), as well as placental surface area (β=-9.6 [-18.3 – −1.0], p=0.03) and largest diameter (β=-0.4 [-0.8, −0.04], p=0.03) compared to controls (Table 4).

**Table 4.**
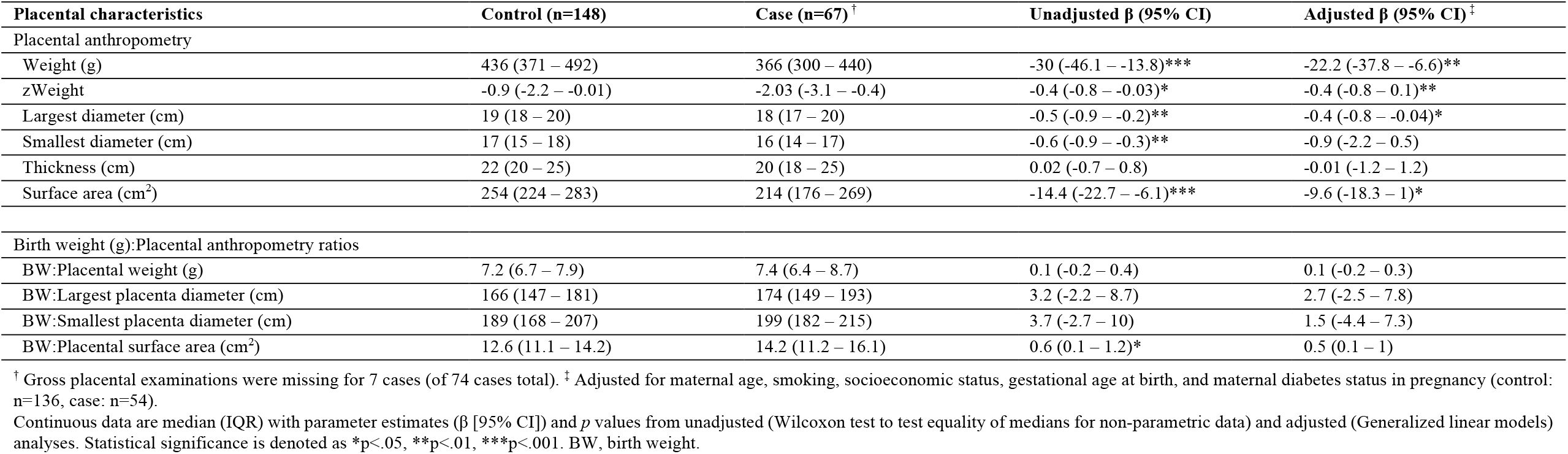
Placental characteristics for infants with isolated neural tube defects and matched controls.

| Placental characteristics | Control (n=148) | Case (n=67) <sup>†</sup> | Unadjusted $\beta$ (95% CI) | Adjusted $\beta$ (95% CI) <sup>‡</sup> |
| --- | --- | --- | --- | --- |
| Placental anthropometry |  |  |  |  |
| Weight (g) | 436 (371 – 492) | 366 (300 – 440) | -30 (-46.1 – -13.8)*** | -22.2 (-37.8 – -6.6)** |
| zWeight | -0.9 (-2.2 – -0.01) | -2.03 (-3.1 – -0.4) | -0.4 (-0.8 – -0.03)* | -0.4 (-0.8 – 0.1)** |
| Largest diameter (cm) | 19 (18 – 20) | 18 (17 – 20) | -0.5 (-0.9 – -0.2)** | -0.4 (-0.8 – -0.04)* |
| Smallest diameter (cm) | 17 (15 – 18) | 16 (14 – 17) | -0.6 (-0.9 – -0.3)** | -0.9 (-2.2 – 0.5) |
| Thickness (cm) | 22 (20 – 25) | 20 (18 – 25) | 0.02 (-0.7 – 0.8) | -0.01 (-1.2 – 1.2) |
| Surface area (cm<sup>2</sup>) | 254 (224 – 283) | 214 (176 – 269) | -14.4 (-22.7 – -6.1)*** | -9.6 (-18.3 – 1)* |
| Birth weight (g):Placental anthropometry ratios |  |  |  |  |
| BW:Placental weight (g) | 7.2 (6.7 – 7.9) | 7.4 (6.4 – 8.7) | 0.1 (-0.2 – 0.4) | 0.1 (-0.2 – 0.3) |
| BW:Largest placenta diameter (cm) | 166 (147 – 181) | 174 (149 – 193) | 3.2 (-2.2 – 8.7) | 2.7 (-2.5 – 7.8) |
| BW:Smallest placenta diameter (cm) | 189 (168 – 207) | 199 (182 – 215) | 3.7 (-2.7 – 10) | 1.5 (-4.4 – 7.3) |
| BW:Placental surface area (cm<sup>2</sup>) | 12.6 (11.1 – 14.2) | 14.2 (11.2 – 16.1) | 0.6 (0.1 – 1.2)* | 0.5 (0.1 – 1) |

<sup>†</sup> Gross placental examinations were missing for 7 cases (of 74 cases total). <sup>‡</sup> Adjusted for maternal age, smoking, socioeconomic status, gestational age at birth, and maternal diabetes status in pregnancy (control: n=136, case: n=54).
Continuous data are median (IQR) with parameter estimates ( $\beta$ [95% CI]) and *p* values from unadjusted (Wilcoxon test to test equality of medians for non-parametric data) and adjusted (Generalized linear models) analyses. Statistical significance is denoted as \**p*<.05, \*\**p*<.01, \*\*\**p*<.001. BW, birth weight.

Overall, cases were more likely to have single umbilical artery (vs. two; aOR=301 [52.6 – 1726], p<.0001) and less likely to have calcification of the maternal placental side (aOR=0.4 [0.2 – 0.7], p=0.002) than controls (Table 5, Figure 2). Odds of having many (vs. few) Hofbauer cells (aOR=3.02 [1.2 – 7.3], p=0.03), stromal fibrosis (aOR=3.0 [1.4 – 6.3], p=0.01) and pathological edema (aOR=3.04 [1.4 – 6.7], p=0.01) in the placental terminal villi were also higher for cases compared to controls (Table 5, Figure 2). Further, case placentae were more likely to be hypermature (aOR=6.8 [3.1 – 14.7], p<.0001) at delivery than controls (Table 5, Figure 2). Lastly, Langhans’ layer was present in eight (10.8%) of case placentae but absent in control placentae, thus, a between-groups comparison was not performed (Table 5, Figure 2).

**Table 5.** Associations between isolated neural tube defects and placental pathologies in cases and controls.

| Placental pathology |  | Control (n=148) | Case (n=74) <sup>†</sup> | Odds of pathology in case placentae (vs. control) |  |
| --- | --- | --- | --- | --- | --- |
|  |  |  |  | OR (95% CI) | aOR (95% CI)<sup>‡</sup> |
| Umbilical cord (n [%]) |  |  |  |  |  |
| Cord edema | Present | 5 (3.4) | 3 (4.1) | 1.4 (0.4 – 4.3) | 0.6 (0.1 – 5.4) |
|  | No description | 142 (96) | 63 (85.1) |  |  |
|  | Unknown | 1 (0.7) | 8 (10.8) |  |  |
| Single umbilical artery | Yes | 1 (0.7) | 6 (8.1) | 14.5 (6.04 – 34.6)*** | 301 (52.6 – 1726)*** |
|  | No | 147 (99.3) | 61 (82.4) |  |  |
|  | Unknown | 0 | 7 (9.5) |  |  |
| Neutrophilic infiltration <sup>‡</sup> | Any | 11 (7.4) | 7 (9.5) | 1.5 (0.70 – 3.4) | 1.8 (0.7 – 4.4) |
|  | Not seen | 137 (92.6) | 57 (77) |  |  |
|  | Unknown | 0 | 10 (13.5) |  |  |
| Membranes & fetal surface (n [%]) |  |  |  |  |  |
| Opacity of membranes | Opaque | 10 (6.8) | 6 (8.1) | 1.4 (0.6 – 3.2) | 1.2 (0.4 – 3.2) |
|  | Not opaque | 138 (96.5) | 59 (79.7) |  |  |
|  | Unknown | 0 | 7 (9.5) |  |  |
| Thrombosed fetal vessels | Any | 0 | 0 | - | - |
|  | Not seen | 148 (100) | 67 (90.5) |  |  |
|  | Unknown | - | 7 (9.5) |  |  |
| Cysts | Any | 9 (6.1) | 3 (4.1) | 0.8 (0.2 – 2.4) | 0.3 (0.1 – 1.2) |
|  | Not seen | 139 (93.9) | 62 (83.7) |  |  |
|  | Unknown | 0 | 9 (12.2) |  |  |
| Neutrophilic infiltration <sup>§</sup> | Any | 28 (18.9) | 13 (17.6) | 1.1 (0.6 – 2) | 1.1 (0.5 – 2.3) |
|  | Not seen | 120 (81.1) | 52 (70.3) |  |  |
|  | Unknown | 0 | 9 (12.2) |  |  |
| Maternal surface (n [%]) |  |  |  |  |  |
| Infarcts | Any | 29 (17.6) | 17 (23) | 1.4 (0.8 – 2.4) | 1.7 (0.9 – 3.1) |
|  | None | 119 (80.4) | 50 (67.6) |  |  |
|  | Unknown | 0 | 7 (9.5) |  |  |
| Calcification | Any | 83 (56.1) | 26 (35.1) | 0.5 (0.3 – 0.8)** | 0.4 (0.2 – 0.7)** |
|  | None | 65 (43.9) | 41 (55.4) |  |  |
|  | Unknown | 0 | 7 (9.5) |  |  |
| Terminal villi (n [%]) |  |  |  |  |  |
| Langhans' layer | Present | 0 | 8 (10.8) | - | - |
|  | Not seen | 148 (100) | 57 (77) |  |  |
|  | Unknown | 0 | 9 (12.2) |  |  |
| Hofbauer cells | Many | 7 (4.7) | 9 (12.2) | 3.2 (1.6 – 6.6)** | 3.02 (1.2 – 7.3)* |
|  | Few | 141 (95.3) | 56 (75.7) |  |  |
|  | Unknown | 0 | 9 (12.2) |  |  |
| Stromal fibrosis | Present | 10 (6.8) | 9 (12.2) | 2.2 (1.1 – 4.5)* | 3 (1.4 – 6.3)* |
|  | Not seen | 138 (93.2) | 56 (75.7) |  |  |
|  | Unknown | 0 | 9 (12.2) |  |  |
| Pathological edema | Present | 12 (8.1) | 11 (14.9) | 2.31 (1.2 – 4.4)* | 3.04 (1.4 – 6.7)* |
|  | Not seen | 136 (91.9) | 54 (73) |  |  |
|  | Unknown | 0 | 9 (12.2) |  |  |
| Apparent maturity of placenta<sup>†</sup> |  |  |  |  |  |
|  | Immature | 34 (23) | 5 (6.8) | 0.3 (0.1 – 0.8)** | 0.01 (0.002 – 0.04)*** |
|  | Hypermaturation | 5 (3.4) | 9 (12.2) | 4.0 (1.9 – 8.2)** | 6.8 (3.1 – 14.7)*** |
|  | Appropriate | 108 (73) | 49 (66.2) | - | - |
|  | Unknown | 1 (0.7) | 11 (14.9) |  |  |

<sup>†</sup> Adjusted for maternal age, smoking, socioeconomic status, gestational age at birth, and maternal diabetes status in pregnancy. <sup>‡</sup> Considers neutrophilic infiltration in the umbilical artery, vein, or cord substance. <sup>§</sup> Considers neutrophilic infiltration amnion and chorion of the membrane and placental surface. <sup>†</sup> Reference group: appropriate maturity.
Data are n (%) and odds ratios or adjusted odds ratios (95% CI) for presence of pathology in case placentae (reference group: controls) from Nominal Logistic regression models with p value from Likelihood Ratio Chi Square test. Statistical significance is denoted as \*p<.05, \*\*p<.01, \*\*\*p<.001. CI, confidence interval. OR, odds ratio. aOR, adjusted odds ratio.

**Figure 2.**
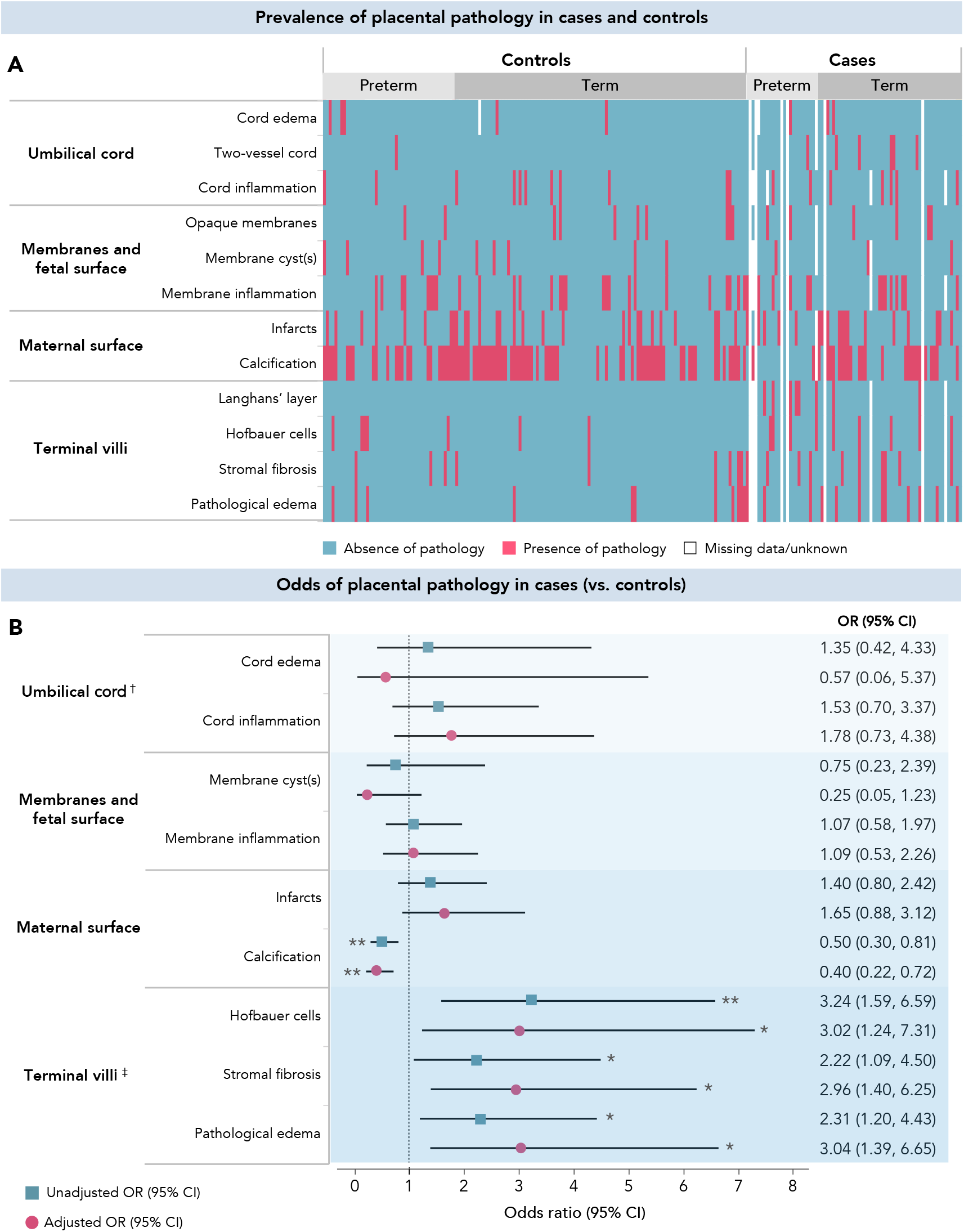
Placental pathology prevalence (A) and odds (B) in cases and controls. (A) Prevalence and absence of placental pathology for case and control infants born preterm (<37 weeks’ gestation, case: n=24, control: n=46) and full term (≥37 weeks’ gestation, case: n=50, control: n=102) are denoted by pink and blue, respectively. White cells represent missing data or data marked as unknown. (B) Odds ratios or adjusted odds ratios (95% CI) for presence of pathology in case placentae (reference group: controls) from Nominal Logistic regression models with p value from Likelihood Ratio Chi Square test (*p<0.05, **p<0.01). The adjusted model includes maternal age, smoking, socioeconomic status, gestational age at delivery, and maternal diabetes status in pregnancy. † Odds of case infants having a single umbilical artery are not included in forest plot due to high range (OR [95% CI]: 14.5 [6.04, 34.6], aOR: 301 [52.6, 1726]). ‡ Odds of cases having Langhans’ layer were not calculated as Langhans’ layer was not seen in any controls. There were 8 cases (10.8%) with Langhans’ layer.

Cases with spina bifida had lower placental surface area (β=-14.4 [-27.9 – −0.8], p=0.04), smallest (β=-0.5 [-1 – −0.03], p=0.04) and largest (β=-0.6 [-1.1 – −0.1], p=0.03) diameters, and higher birthweight-to-placental weight (β=0.4 [0.1 – 0.7], p=0.02), surface area (β=1.0 [0.3 – 1.6], p=0.005), and largest (β=7.2 [1.3 –13.2], p=0.02) and smallest diameter (β=7.5 [0.2 – 14.8], p=0.04) ratios compared to controls (Table 6). Cases with spina bifida also had increased odds of neutrophilic infiltration in the umbilical cord (aOR=15.6 [4.2 – 58.1], p<.001) and fetal membranes (aOR=3.7 [1.3 – 10.8], p=0.02), membrane cysts (aOR=25.6 [4 – 164], p=0.001) and stromal fibrosis (aOR=3.9 [1.4 – 10.8], p=0.009; Table 7).

**Table 6.** Placental characteristics for cases with spina bifida or anencephaly in comparison to matched controls.

| Placental characteristics | Control<br>(n=66) | Spina bifida<br>(n=31) † | Unadjusted<br>β (95% CI) | Adjusted<br>β (95% CI) † | Control<br>(n=66) | Anencephaly<br>(n=29) † | Unadjusted<br>β (95% CI) | Adjusted<br>β (95% CI) † |
| --- | --- | --- | --- | --- | --- | --- | --- | --- |
| Gross placental anthropometry |  |  |  |  |  |  |  |  |
| Weight (g) | 432 (353 – 489) | 360 (300 – 460) | -27.9 (-55.2 – -0.6)* | -18.5 (-300 – 168) | 440 (379 – 483) | 335 (271 – 400) | -42.5 (-62.5 – -22.6)*** | -37.8 (-59.8 – -15.8)<br>** |
| zWeight | -0.9 (-2.2 – 0.02) | -1.9 (-3.7 – 0.1) | -0.3 (-0.9 – 0.3) | -0.4 (-0.9 – 0.2) | -0.8 (-1.8 – -0.1) | -2.4 (-3.04 – -1.5) | -0.6 (-1.1 – -0.1)** | -0.7 (-1.2 – 0.2)* |
| Largest diameter (cm) | 19 (18 – 20) | 17 (16 – 19) | -0.8 (-1.3 – -0.2)** | -0.6 (-1.1 – -0.1)* | 20 (18 – 21) | 19 (17 – 21) | -0.3 (-0.8 – 0.2) | 0.03 (-0.5 – 0.6) |
| Smallest diameter (cm) | 17 (15 – 18) | 16 (14 – 17) | -0.7 (-1.2 – -0.2)* | -0.5 (-1 – -0.03)* | 17 (15.5 – 18) | 15 (14 – 17) | -0.6 (-1.04 – -0.2)* | -0.4 (-0.8 – 0.1) |
| Thickness (cm) | 20 (19 – 25) | 21 (17 – 25) | 0.1 (-1.1 – 1.3) | 0.3 (-0.9 – 1.5) | 23 (20 – 25) | 20 (18.5 – 25) | -0.3 (-1.4 – 0.8) | -0.2 (-1.4 – 1.1) |
| Surface area (cm<sup>2</sup>) | 254 (212 – 286) | 214 (176 – 240) | -19.2 (-33 – -5.5)** | -14.4 (-27.9 – -0.8)* | 254 (224 – 288) | 214 (180 – 280) | -10.8 (-22.2 – 0.7)* | -3 (-15.4 – 9.4) |
| BW:Placental anthropometry ratios |  |  |  |  |  |  |  |  |
| BW:Placental weight | 7.2 (6.5 – 8.1) | 7.8 (6.7 – 9.2) | 0.5 (0.1 – 0.9) | 0.4 (0.1 – 0.7)* | 7.2 (6.7 – 7.8) | 7.3 (4.8 – 8.5) | -0.3 (-0.7 – 0.2) | -0.6 (-1.1 – -0.1)* |
| BW:Largest placenta diameter | 166 (145 – 184) | 178 (163 – 201) | 10.2 (2.7 – 17.6)* | 7.2 (1.3 – 13.2)* | 166 (147 – 180) | 132 (106 – 169) | -14.3 (-23.3 – -5.4)* | -22.3 (-32.5 – -12)<br>*** |
| BW:Smallest placenta diameter | 184 (162 – 215) | 201 (188 – 225) | 10.9 (2.3 – 19.6)* | 7.5 (0.2 – 14.8)* | 191 (172 – 206) | 183 (118 – 212) | -11.9 (-22.6 – -1.3) | -20.6 (-31.9 – -9.3)<br>*** |
| BW:Placental surface area | 12.6 (10.5 – 14.2) | 14.9 (12.8 – 16.1) | 1.2 (0.4 – 1.9)** | 1.0 (0.3 – 1.6)** | 12.4 (11.3 – 13.5) | 10.1 (8.3 – 15.8) | -0.7 (-1.6 – 0.2) | -1.8 (-2.8 – -0.7)** |

† Gross placental examinations were missing for 2 cases with spina bifida (of 33 total) and 4 cases with anencephaly (of 33 total). ‡ Adjusted for maternal age, smoking, socioeconomic status, gestational age at birth, and maternal diabetes status in pregnancy.
Continuous data are median (IQR) with parameter estimates (β [95% CI]) and *p* values from unadjusted (Wilcoxon test to test equality of medians for non-parametric data) and adjusted (Generalized linear models) analyses. Statistical significance is denoted as \**p*<.05, \*\**p*<.01, \*\*\**p*<.001. BW, birth weight.

**Table 7.** Odds of placental pathology for infants with spina bifida or anencephaly in comparison to matched controls.

| Placental pathology |  | Odds of placental pathology in cases with spina bifida |  |  |  | Odds of placental pathology in cases with anencephaly |  |  |  |
| --- | --- | --- | --- | --- | --- | --- | --- | --- | --- |
|  |  | Control (n=66) | Spina bifida (n=33) | OR (95% CI) | aOR (95% CI) † | Control (n=66) | Anencephaly (n=33) | OR (95% CI) | aOR (95% CI) † |
| Umbilical cord (n [%]) |  |  |  |  |  |  |  |  |  |
| Cord edema | Present | 0 | 1 (3.03) | - | - | 5 (7.6) | 2 (6.1) | 0.9 (0.2 – 3.9) | 1.1 (0.1 – 9.2) |
|  | No description | 66 (100) | 30 (90.9) |  |  | 60 (90.9) | 26 (78.8) |  |  |
|  | Unknown | 0 | 2 (6.1) |  |  | 1 (1.5) | 5 (15.2) |  |  |
| Single umbilical artery | Yes | 0 | 3 (9.1) | - | - | 1 (1.5) | 3 (9.1) | 7.5 (2.2 – 25.4)** | 2278 (194 – 26824)*** |
|  | No | 66 (100) | 28 (84.9) |  |  | 65 (98.5) | 26 (78.8) |  |  |
|  | Unknown | 0 | 2 (6.1) |  |  | 0 | 4 (12.1) |  |  |
| Neutrophilic infiltration† | Any | 6 (9.1) | 7 (21.2) | 3.04 (1.3 – 7.1)* | 15.6 (4.2 – 58.1)*** | 4 (6.1) | 0 | - | - |
|  | Not seen | 60 (90.9) | 23 (69.7) |  |  | 62 (93.9) | 27 (81.8) |  |  |
|  | Unknown | 0 | 3 (9.1) |  |  | 0 | 6 (18.2) |  |  |
| Membranes & fetal surface (n [%]) |  |  |  |  |  |  |  |  |  |
| Thrombosed fetal vessels | Any | 0 | 0 | - | - | 0 | 0 | - | - |
|  | Not seen | 66 (100) | 31 (93.9) |  |  | 66 (100) | 29 (87.9) |  |  |
|  | Unknown | 0 | 2 (6.1) |  |  | 0 | 4 (12.1) |  |  |
| Cysts | Any | 1 (1.5) | 3 (9.1) | 7.0 (2.1 – 23.5)** | 25.6 (4 – 164)*** | 8 (12.1) | 0 | - | - |
|  | Not seen | 65 (98.5) | 28 (84.9) |  |  | 58 (87.9) | 27 (81.8) |  |  |
|  | Unknown | 0 | 2 (6.1) |  |  | 0 | 6 (18.2) |  |  |
| Neutrophilic infiltration§ | Any | 12 (18.2) | 9 (27.3) | 1.8 (0.8 – 4.01) | 3.7 (1.3 – 10.8)* | 13 (19.7) | 4 (12.1) | 0.7 (0.3 – 2.1) | 0.7 (0.2 – 3.0) |
|  | Not seen | 54 (81.8) | 22 (66.7) |  |  | 53 (80.3) | 23 (69.7) |  |  |
|  | Unknown | 0 | 1 (3.03) |  |  | 0 | 6 (18.2) |  |  |
| Maternal surface (n [%]) |  |  |  |  |  |  |  |  |  |
| Infarcts | Any | 14 (21.2) | 2 (6.1) | 0.3 (0.1 – 1.1) | 0.3 (0.1 – 1.1) | 10 (15.2) | 11 (33.3) | 3.4 (1.6 – 7.3)** | 5.8 (2.2 – 15)*** |
|  | None | 52 (78.8) | 29 (87.9) |  |  | 56 (84.9) | 18 (54.6) |  |  |
|  | Unknown | 0 | 2 (6.1) |  |  | 0 | 4 (12.1) |  |  |
| Calcification | Any | 30 (45.5) | 11 (33.3) | 0.7 (0.3 – 1.4) | 0.8 (0.3 – 1.8) | 43 (65.2) | 10 (30.3) | 0.3 (0.1 – 0.6)** | 0.2 (0.1 – 0.4)*** |
|  | None | 36 (54.6) | 20 (60.6) |  |  | 23 (34.9) | 19 (57.6) |  |  |
|  | Unknown | 0 | 2 (6.1) |  |  | 0 | 4 (12.1) |  |  |
| Terminal villi (n [%]) |  |  |  |  |  |  |  |  |  |
| Langhans' layer | Present | 0 | 4 (12.1) | - | - | 0 | 4 (12.1) | - | - |
|  | Not seen | 66 (100) | 27 (81.8) |  |  | 66 (100) | 23 (69.7) |  |  |
|  | Unknown | 0 | 2 (6.1) |  |  | 0 | 6 (18.2) |  |  |
| Hofbauer cells | Many | 5 (7.6) | 6 (18.2) | 2.9 (1.2 – 7.2)* | 1.6 (0.4 – 6.2) | 2 (3.03) | 2 (6.1) | 2.6 (0.6 – 10.9) | 5.01 (1.02 – 24.7)* |
|  | Few | 61 (92.4) | 25 (75.8) |  |  | 64 (97.0) | 25 (75.8) |  |  |
|  | Unknown | 0 | 2 (6.1) |  |  | 0 | 6 (18.2) |  |  |
| Stromal fibrosis | Present | 5 (7.6) | 7 (21.2) | 3.6 (1.5 – 8.3)** | 3.9 (1.4 – 10.8)** | 5 (7.6) | 2 (6.1) | 1.0 (0.2 – 4.1) | 2.4 (0.5 – 10.7) |
|  | Not seen | 61 (92.4) | 24 (72.7) |  |  | 61 (92.4) | 25 (75.8) |  |  |
|  | Unknown | 0 | 2 (6.1) |  |  | 0 | 6 (18.2) |  |  |
| Pathological edema | Present | 7 (10.6) | 4 (12.1) | 1.3 (0.4 – 3.6) | 1.01 (0.2 – 4.4) | 5 (7.6) | 6 (18.2) | 3.5 (1.4 – 8.7)** | 5.0 (1.5 – 16.1)** |
|  | Not seen | 59 (89.4) | 27 (81.8) |  |  | 61 (92.4) | 21 (63.6) |  |  |
|  | Unknown | 0 | 2 (6.1) |  |  | 0 | 6 (18.2) |  |  |
| Apparent maturity of placenta<sup>1</sup> |  |  |  |  |  |  |  |  |  |
|  | Immature | 10 (15.2) | 3 (9.1) | 0.7 (0.2 – 2.4) | 0.1 (0.01 – 0.9)* | 20 (30.3) | 2 (6.1) | 0.2 (0.1 – 1.0)* | 0.002 (0.0003 – 0.02)*** |
|  | Hypermaturation | 2 (3.03) | 3 (9.1) | 3.5 (1.04 – 11.9)* | 6.31 (1.6 – 25.3)** | 3 (4.6) | 6 (18.2) | 4.4 (1.8 – 11.2)** | 8.6 (2.7 – 26.7)*** |
|  | Appropriate | 54 (81.8) | 23 (69.7) |  |  | 42 (63.6) | 19 (57.6) |  |  |
|  | Unknown | 0 | 4 (12.1) |  |  | 0 | 6 (18.2) |  |  |

† Adjusted for maternal age, smoking, socioeconomic status, gestational age at birth, and maternal diabetes status in pregnancy. ‡ Considers neutrophilic infiltration in the umbilical artery, vein, or cord substance. § Considers neutrophilic infiltration amnion and chorion of the membrane and placental surface. <sup>1</sup> Reference group: appropriate maturity.
Data are n (%) and odds ratios or adjusted odds ratios (95% CI) for presence of pathology in case placentae (reference group: controls) from Nominal Logistic regression models with p value from Likelihood Ratio Chi Square test. Statistical significance is denoted as \*p<.05, \*\*p<.01, \*\*\*p<.001. CI, confidence interval. OR, odds ratio. aOR, adjusted odds ratio.

Cases with anencephaly had lower placental weight (β=-37.8 [-59.8 – −15.8], p=0.001), gestational-age standardized placental weight (β=-0.7 [-1.2 – 0.2], p=0.01), and lower birthweight-to-placental surface area (β=-1.8 [-2.8 – −0.7], p=0.001), weight (β=-0.6 [-1.1 – −0.1], p=0.02), and smallest (β=-20.6 [-31.9 – - 9.3], p=0.001) and largest diameter (β=-22.3 [-32.5 – - 12], p<.0001) ratios compared to controls (Table 6). Cases with anencephaly also had increased odds of having a single umbilical artery (aOR=2278 [194 – 26824], p<.0001), maternal surface infarcts (aOR=5.8 [2.2 – 15], p=0.0003), many (vs. few) Hofbauer cells (aOR=5.01 [1.02 – 24.7], p=0.05) and pathological edema (aOR=5.0 [1.5 – 16.1], p=0.008; Table 7). Odds of placental hypermaturity were increased in cases with spina bifida (aOR=6.31 [1.6 – 25.3], p=0.03) and anencephaly (8.6 [2.7 – 26.7], p=0.0002; Table 7). There were no differences in birth outcomes for placental characteristics or pathology for cases with encephalocele compared to matched controls (Supplementary Tables 2 and 3).

In subgroup analysis of preterm (Supplementary Table 4) and term (Supplementary Table 5) placentae, preterm cases had lower placental weight (β=-48.4 [-74.5 – - 22.2], p=0.001), and lower birthweight-to-largest (β=-11.8 [-23.2 – −0.45], p=0.04) and smallest diameter (β=-15.0 [-28.8 – −1.10], p=0.04) ratios compared to preterm controls. Odds of maternal surface calcification (aOR=0.03 [0.01 – 0.2], p<.0001), placental immaturity (aOR=0.05 [0.01, 0.22], p<.0001), and membrane cysts (aOR=0.03 [0.001 – 0.50], p=0.02) were lower in preterm cases than preterm controls. Term cases had higher birthweight-to-placental surface area ratio (β=0.71 [0.17 – 1.24], p=0.01) and were more likely to have many (vs. few) Hofbauer cells (aOR=7.78 [2.46 – 24.6], p<.001), stromal fibrosis (aOR=2.98 [1.18 – 7.53], p=0.03), pathological edema (aOR=3.62 [1.48 – 8.82], p=0.005) and placental hypermaturity (aOR=5.73 [2.40 – 13.7], p<.0001) than term controls.

### Sex-based differences in placental characteristics

Placental hypermaturity was more prevalent in female than male cases (OR=6.52 [1.08 – 126], p=0.04; Supplementary Table 6, Figure 3). All female cases with placental hypermaturity were born full term (Figure 3). No other sex-based differences were identified.

**Figure 3.**
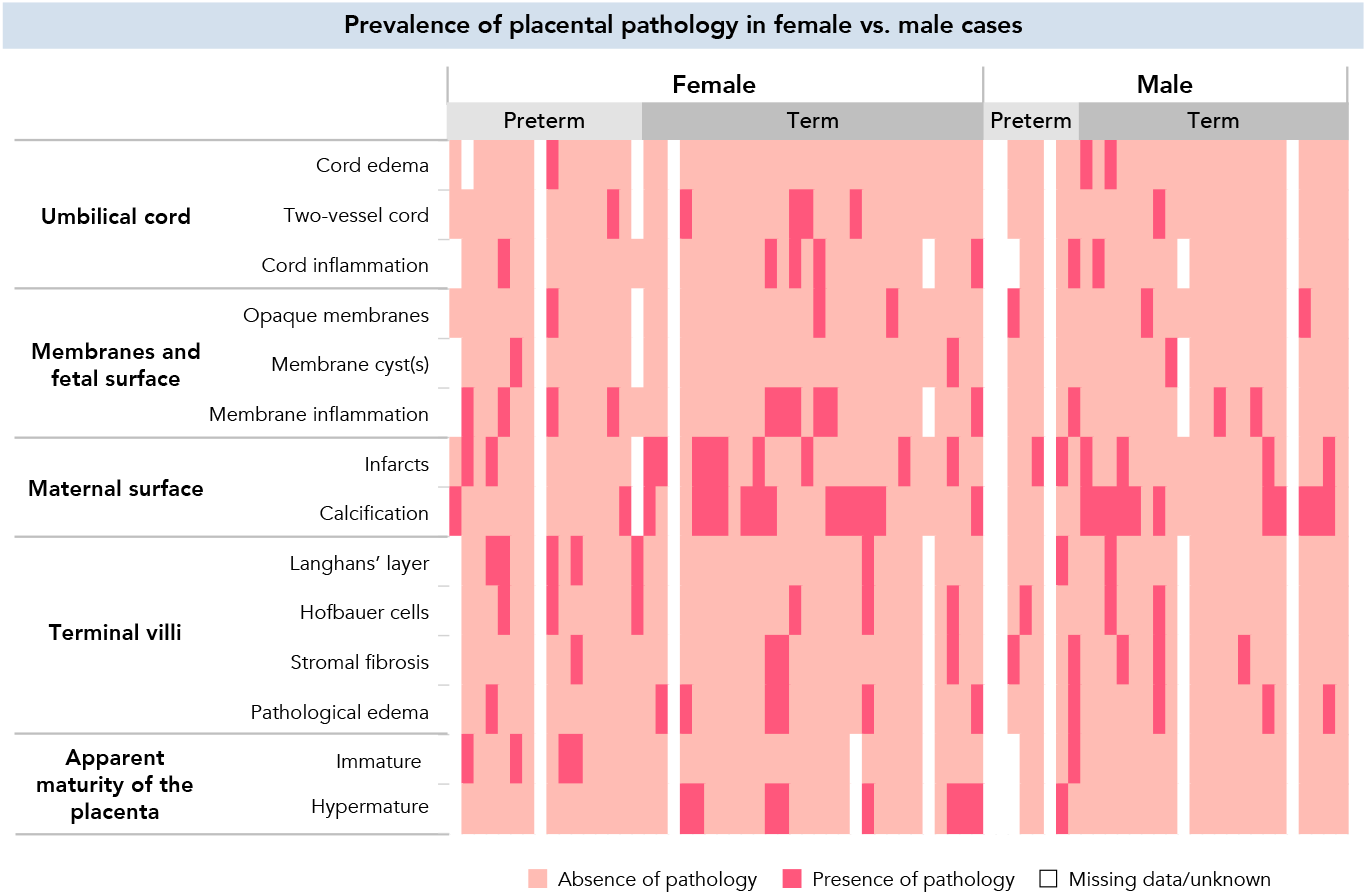
Placental pathology prevalence in cases stratified by sex. Presence (dark pink) and absence (light pink) of placental pathology for female and male cases born preterm (<37 weeks’ gestation; female: n=16, male: n=8) and full term (≥37 weeks’ gestation; female: n=28, male: n=22). White cells represent missing data or data marked as unknown.

## Discussion

Using data from the CPP and a case-cohort study design, we found that overall, fetuses with NTDs had higher risk of pathology in the umbilical cord and placenta and lower birth length compared to controls. Both cases with spina bifida and anencephaly were smaller at birth and had placental hypermaturity, while umbilical cord and fetal membrane inflammation was increased in cases with spina bifida, specifically. These findings suggest that fetuses with isolated NTDs are at an increased risk of altered placental development and suboptimal growth outcomes in comparison to a well-matched control cohort.

Cases with spina bifida experienced a higher risk of placental hypermaturity and higher birthweight-to placental size ratios compared to controls. Accelerated placental maturation, demonstrated by adaptations in the placental villi, may be a compensatory response to meet fetal requirements, and has been associated with improved neonatal outcomes in infants born preterm in the CPP cohort^32^. Notably, pathology in the terminal villi and accelerated placental maturation were mainly present in term cases, and all term cases with placental hypermaturity were female. This aligns with previous consensus that female placentae are more likely than males to exhibit multiple adaptations in high-risk pregnancies in an aim to preserve fetal survival^19^. This adaptation may have contributed to the higher fetal weight-to-placental size ratios we observed in cases with spina bifida. However, cases with spina bifida were still more likely than matched controls to be born extremely preterm and with a lower body length, and have increased neutrophilic infiltration in the umbilical cord and fetal membranes, which is characteristic of ascending fetal inflammation and associates with preterm birth and increased perinatal morbidity^33^. Together, these findings may suggest that placental maladaptations in pregnancies with isolated fetal spina bifida may contribute to increased morbidity in these offspring.

While cases with anencephaly also had an increased risk of placental hypermaturity and terminal villi pathology, they had lower birthweight-to-placental size ratios than controls, which may be indicative of reduced placental efficiency^34^ and may compromise maternal-fetal exchange^35^. These cases also had increased odds of having a single umbilical artery, which is consistent with reports of population-level associations between congenital anomalies of the brain and spinal cord and single umbilical artery^36^ and is associated with low birthweight^37^. Together, these pathologies may have led to the lower birthweight and younger gestational age at birth we observed in cases with anencephaly in comparison to controls.

Cases were also more likely to have pathological edema and stromal fibrosis in the terminal villi than controls, which both associate with intrauterine growth restriction^38,39^. Hofbauer cells are important regulators of placental vasculogenesis^40^ and inflammation, may promote fibrosis in the placental villi^41^, and were also elevated in cases compared to controls. To our knowledge, neither dysregulation of Hofbauer cells, nor edema or stromal fibrosis have previously been reported in placentae from fetuses with NTDs. Placental Hofbauer cells and monocytes are folate-dependent and express folate receptor-beta (FR-β)^42^. Thus, it is plausible that altered folate bioavailability in pregnancies with isolated fetal NTDs may lead to Hofbauer cell dysregulation and subsequently altered vasculogenesis and stromal fibrosis, as we report here. Reduced levels of Hofbauer cells with an anti-inflammatory phenotype and decreased FR-β mRNA and protein expression by Hofbauer cells have been reported in placentae from pregnancies with severe pre-eclampsia^43^. These changes are hypothesized to contribute to the anti-angiogenic environment and placental structural abnormalities often recorded in placentae from preeclamptic pregnancies^43^, suggesting a possible relationship between folate uptake by Hofbauer cells, Hofbauer cell activity and placental development. How reduced folate bioavailability may contribute to Hofbauer cell dysregulation, including shifts between pro- and anti-inflammatory phenotypes, and whether this may lead to placental pathology should be further explored in models of isolated NTDs where causality can be better scrutinized.

That peri-conceptional folic acid supplementation and fortification of wheat products with folic acid were not introduced until well after the CPP concluded, is a key limitation to our study. Yet, our findings are relevant where adherence to folic acid supplementation is poor, or in countries where there is no universal folic acid fortification (nearly 60% of countries worldwide in 2017^44^). Additionally, altered DNA methylation in the chorionic villi of placentae from a folate-replete population suggests that pregnancies with fetal NTDs may remain at risk of altered placental development^49^, even when maternal folate status is sufficient. Lastly, an estimated one in five non-pregnant women in the United States may have blood folate levels below those recommended for NTD prevention^50^, and the growing number of people who cannot consume wheat products^51^ may also be at higher risk of folate deficiency^52^, suggesting that our findings from a cohort that likely experienced higher rates of folate deficiency may still be relevant for some pregnancies with fetal NTDs today.

Other potential limitations to our study include population-level changes that have occurred since the CPP concluded, including the increased prevalence of maternal pre-pregnancy overweight or obesity. Approximately 24.8% of the mothers in our case-cohort study had a pre-pregnancy BMI classified as overweight or obese, while more recent estimates report that in the United States, 51.4% of people enter pregnancy with overweight or obesity^53^. High maternal pre-pregnancy BMI is a known risk factor for fetal NTDs^6^ and associates with altered placental phenotype^18^, and would itself be a critical exposure of interest in a present day cohort. Further, Hispanic women in this United States experience higher rates spina bifida and anencephaly than non-Hispanic White and non-Hispanic Black women, and were underrepresented in this cohort^54^. Genetic testing data were also not available for prenatal diagnoses of NTDs in these cases, so it is plausible that the true incidence of NTDs was not captured in this cohort. Lastly, as placental dysfunction has been causally linked to preterm birth^11^, the controls in our cohort who were born preterm may be at an increased risk of suboptimal placental development themselves, which may have confounded our comparisons.

Strengths of our study include the use of data from the CPP, which remains the largest prospective, multicentre birth cohort in the United States, our strict selection for cases with isolated NTDs, and matching criteria for identification of a control cohort. Further, the data examined here would be challenging to collect in contemporary cohorts for such a large number of cases due to advances in prenatal diagnostics and increased pregnancy termination of pregnancies complicated by fetal NTDs.

In summary, our study comprehensively assesses the prevalence of placental pathology in a large cohort of fetuses with isolated NTDs, and increases our understanding of the mechanisms that may drive poor fetal growth and preterm birth in these pregnancies. Future studies should evaluate the potential benefits of interventions for the prevention of NTDs on placental development and function. Understanding the mechanisms that may contribute to the risk of mortality or subsequent morbidities in fetuses with isolated NTDs is critical for determining how to improve survival and developmental trajectories in these infants.

## Data Availability

The data that support the findings of this study are available in the United States National Archives at https://www.archives.gov/research/electronic-records/nih.html, National Archives Identifier: 606622 (Record Group 443: Records of the National Institutes of Health [NIH]). Derived data supporting the findings of this study are available from the corresponding author on request.

## Acknowledgments

The authors thank Molly Campbell for her assistance with data mining.

## Supplementary tables

**Supplementary Table 1.**
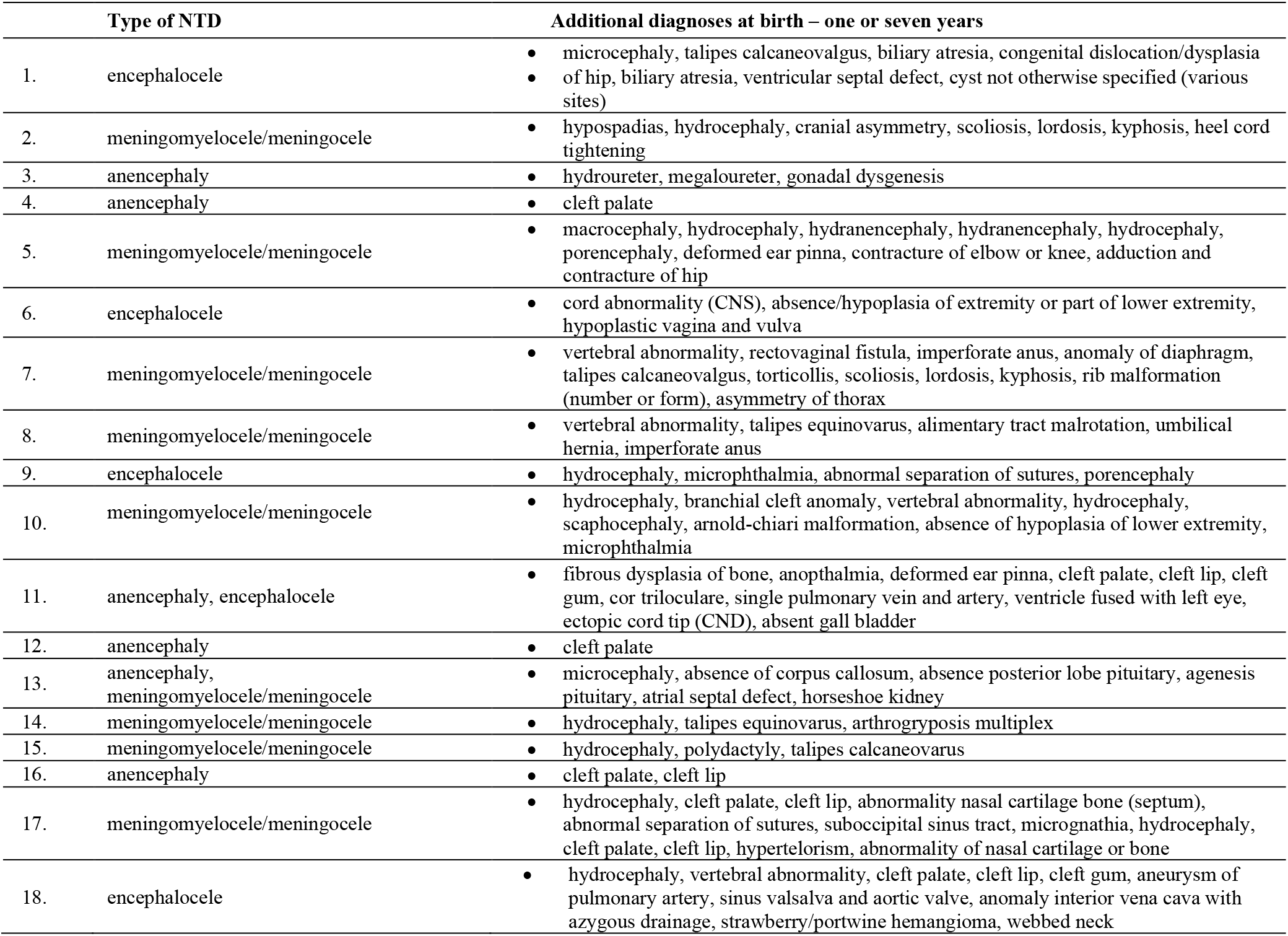
Co-morbid diagnoses of fetuses with a neural tube defect whom were excluded during case identification, Collaborative Perinatal Project (1959-1974).

**Supplementary Table 2.** Infant birth outcomes for cases with encephalocele in comparison to matched controls.

| Infant characteristics | Control (n=16) | Encephalocele (n=8)<sup>†</sup> | Unadjusted $\beta$ (95% CI) | Adjusted $\beta$ (95% CI)<sup>‡</sup> |
| --- | --- | --- | --- | --- |
| Male-to-female sex ratio | 1.67 | 1.67 | - | - |
| GA at birth (weeks)<sup>§</sup> | 40.0 (33.0 – 41.0) | 38.0 (32.8 – 40.3) | -0.63 (-3.63 – 2.36) | 0.67 (-1.44 – 2.79) |
| <37 weeks GA at birth (n [%]) | 4 (25.0) | 2 (25.0) |  |  |
| Sub-categories of PTB (n [%]) |  |  | 0 (-1.5 – 1.5)<sup>¶</sup> |  |
| Moderate/late preterm | 2 (12.5) | 1 (12.5) |  |  |
| Very preterm | 0 | 0 |  |  |
| Extremely preterm | 2 (12.5) | 1 (12.5) |  |  |
| zWeight at birth | -0.07 (-0.71 – 0.88) | 0.15 (-0.18 – 0.94) | 0.001 (-0.76 – 0.76) | -0.15 (-1.03 – 0.72) |
| zLength at birth | 0.82 (-0.15 – 1.54) | 0.67 (-0.67 – 2.16) | -0.007 (-0.8 – 0.79) | 0.36 (-1.23 – 0.50) |
| zHead circumference at birth | 0.22 (-0.71 – 1.12) | 2.37 (0.34 – 5.35) | 1.16 (0.12 – 2.21) | 0.56 (-0.64 – 1.75) |
| Apgar at one minute (/10) | 9.00 (7.00 – 9.00) | 7.00 (2.00 – 8.00) | -1.50 (-2.55 – -0.43)* | -0.94 (-2.52 – 0.64) |
| Apgar at five minutes (/10) | 9.00 (9.00 – 10.0) | 8.00 (5.00 – 9.00) | -1.07 (-1.64 – -0.49)** | -0.31 (-1.10 – 0.48) |
| **Placental characteristics** |  |  |  |  |
| Gross placental anthropometry |  |  |  |  |
| Weight (g) | 429 (330 – 514) | 485 (390 – 550) | 12.6 (-46.0 – 71.2) | -1.39 (-62.2 – 59.4) |
| zWeight | -2.15 (-3.01 – 0.3) | -0.86 (-2.89 – 0.47) | 0.18 (-1.12 – 1.49) | 0.07 (-1.61 – 1.74) |
| Largest diameter (cm) | 20.0 (17.0 – 21.5) | 19.0 (15.0 – 20.0) | -0.66 (-1.85 – 0.54) | -0.31 (-1.41 – 0.79) |
| Smallest diameter (cm) | 16.0 (15.0 – 17.0) | 16.0 (14.0 – 19.0) | -0.13 (-1.09 – 0.83) | -0.30 (-1.09 – 0.49) |
| Thickness (cm) | 22.0 (18.0 – 25.0) | 25.0 (18.0 – 27.0) | 0.98 (-1.47 – 3.42) | 0.89 (-1.75 – 3.52) |
| Surface area (cm<sup>2</sup>) | 239 (214 – 271) | 214 (165 – 299) | -8.08 (-33.8 – 17.8) | -6.42 (-27.9 – 15.0) |
| BW:Placental anthropometry ratios |  |  |  |  |
| BW:Placental weight | 7.31 (6.71 – 8.46) | 6.44 (5.38 – 7.83) | -0.64 (-1.62 – 0.34) | -1.02 (-2.42 – 0.37) |
| BW:Largest placenta diameter | 159 (149 – 178) | 187 (130 – 202) | -1.60 (-14.9 – 18.1) | -7.39 (-27.4 – 12.6) |
| BW:Smallest placenta diameter | 197 (179 – 202) | 199 (137 – 255) | -0.68 (-23.9 – 22.5) | -7.16 (-32.9 – 18.6) |
| BW:Placental surface area | 13.0 (11.1 – 14.6) | 12.3 (8.74 – 19.8) | 0.32 (-1.37 – 2.02) | -0.34 (-2.22 – 1.54) |

<sup>†</sup> Gross placental examinations were missing for 2 cases with encephalocele (of 8 total). <sup>‡</sup> Adjusted for maternal age, smoking, socioeconomic status, gestational age at birth, and maternal diabetes status in pregnancy. For analyses of infant birth anthropometry (standardized for gestational age- and sex), gestational age at birth or distribution of sub-categories of preterm birth, gestational age at birth was not included. <sup>§</sup> Gestational age at birth was marked as missing for infants with a gestational age that was >43 weeks (cases with encephalocele: n=2, matched-controls: n=2). <sup>||</sup> Moderate/late preterm: 32-36 weeks' gestation, very preterm: 28-32 weeks' gestation, extremely preterm: <28 weeks' gestation. <sup>¶</sup> Effect estimate is Kendall's $\tau$ (95% CI).
Continuous data are median (IQR) with parameter estimates ( $\beta$ [95% CI]) and $p$ values from unadjusted (Wilcoxon test to test equality of medians) and adjusted (Generalized linear models) analyses. Categorical data are n (%) with Kendall's $\tau$ (95% CI) and $p$ values from Nominal Logistic regression models (Likelihood Ratio Chi Square test to test associations between study groups and variable distributions). Statistical significance is denoted as \* $p$ <.05, \*\* $p$ <.01, \*\*\* $p$ <.001. GA, gestational age. PTB, preterm birth. BW, birth weight.

**Supplementary Table 3.** Odds of placental pathology for infants with encephalocele in comparison to matched controls.

| Placental pathology |  | Control (n=16) | Case (n=8) | Odds of pathology in encephalocele placentae (vs. control) |  |
| --- | --- | --- | --- | --- | --- |
|  |  |  |  | OR (95% CI) | aOR (95% CI)<sup>†</sup> |
| Umbilical cord (n [%]) |  |  |  |  |  |
| Cord edema | Present | 0 | 0 | - | - |
|  | No description | 16 (100) | 7 (87.5) |  |  |
|  | Unknown | 0 | 1 (12.5) |  |  |
| Single umbilical artery | Yes | 0 | 0 | - | - |
|  | No | 16 (100) | 7 (87.5) |  |  |
|  | Unknown | 0 | 1 (12.5) |  |  |
| Neutrophilic infiltration<sup>‡</sup> | Any | 1 (6.25) | 0 | - | - |
|  | Not seen | 15 (93.8) | 7 (87.5) |  |  |
|  | Unknown | 0 | 1 (12.5) |  |  |
| Membranes & fetal surface (n [%]) |  |  |  |  |  |
| Thrombosed fetal vessels | Any | 0 | 0 | - | - |
|  | Not seen | 16 (100) | 7 (87.5) |  |  |
|  | Unknown | 0 | 1 (12.5) |  |  |
| Cysts | Any | 0 | 0 | - | - |
|  | Not seen | 16 (100) | 7 (87.5) |  |  |
|  | Unknown | 0 | 1 (12.5) |  |  |
| Neutrophilic infiltration<sup>§</sup> | Any | 3 (18.8) | 0 | - | - |
|  | Not seen | 16 (100) | 7 (87.5) |  |  |
|  | Unknown | 0 | 1 (12.5) |  |  |
| Maternal surface (n [%]) |  |  |  |  |  |
| Infarcts | Any | 5 (31.3) | 4 (50.0) | 2.93 (0.65 – 13.2) | - |
|  | None | 11 (68.8) | 3 (37.5) |  |  |
|  | Unknown | 0 | 1 (12.5) |  |  |
| Calcification | Any | 10 (62.5) | 5 (62.5) | 1.50 (0.29 – 7.77) | 0.18 (0.01 – 4.02) |
|  | None | 6 (37.5) | 2 (25.0) |  |  |
|  | Unknown | 0 | 1 (12.5) |  |  |
| Terminal villi (n [%]) |  |  |  |  |  |
| Langhans' layer | Present | 0 | 0 | - | - |
|  | Not seen | 16 (100) | 7 (87.5) |  |  |
|  | Unknown | 0 | 1 (12.5) |  |  |
| Hofbauer cells | Many | 0 | 1 (12.5) | - | - |
|  | Few | 16 (100) | 6 (75.0) |  |  |
|  | Unknown | 0 | 1 (12.5) |  |  |
| Stromal fibrosis | Present | 0 | 0 | - | - |
|  | Not seen | 16 (100) | 7 (87.5) |  |  |
|  | Unknown | 0 | 1 (12.5) |  |  |
| Pathological edema | Present | 0 | 1 (12.5) | - | - |
|  | Not seen | 16 (100) | 6 (75.0) |  |  |
|  | Unknown | 0 | 1 (12.5) |  |  |
| Apparent maturity of placenta<sup>†</sup> |  |  |  |  |  |
|  | Immature | 4 (25.0) | 0 | - | - |
|  | Hypermaturation | 0 | 0 | - | - |
|  | Appropriate | 12 (75.0) | 7 (87.5) |  |  |
|  | Unknown | 0 | 1 (12.5) |  |  |

<sup>†</sup> Adjusted for maternal age, smoking, socioeconomic status, gestational age at birth, and maternal diabetes status in pregnancy. <sup>‡</sup> Considers neutrophilic infiltration in the umbilical artery, vein, or cord substance. <sup>§</sup> Considers neutrophilic infiltration amnion and chorion of the membrane and placental surface. <sup>†</sup> Reference group: appropriate maturity.
Data are n (%) and odds ratios and adjusted odds ratios (95% CI) for presence of pathology in case placentae (reference group: controls) from Nominal Logistic regression models with p value from Likelihood Ratio Chi Square test. Statistical significance is denoted as \*p<.05, \*\*p<.01, \*\*\*p<.001. CI, confidence interval. OR, odds ratio. aOR, adjusted odds ratio.

**Supplementary Table 4.**
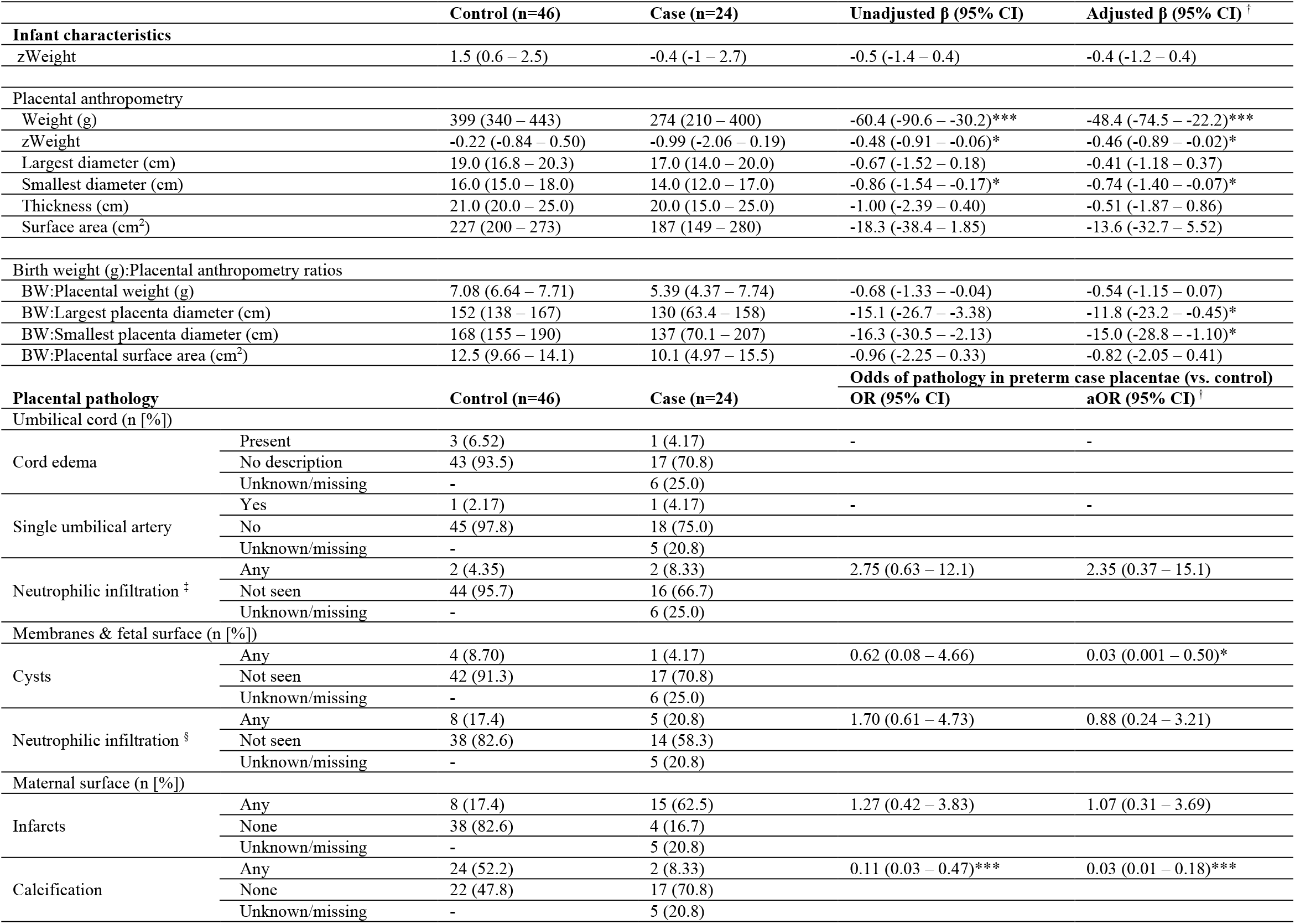
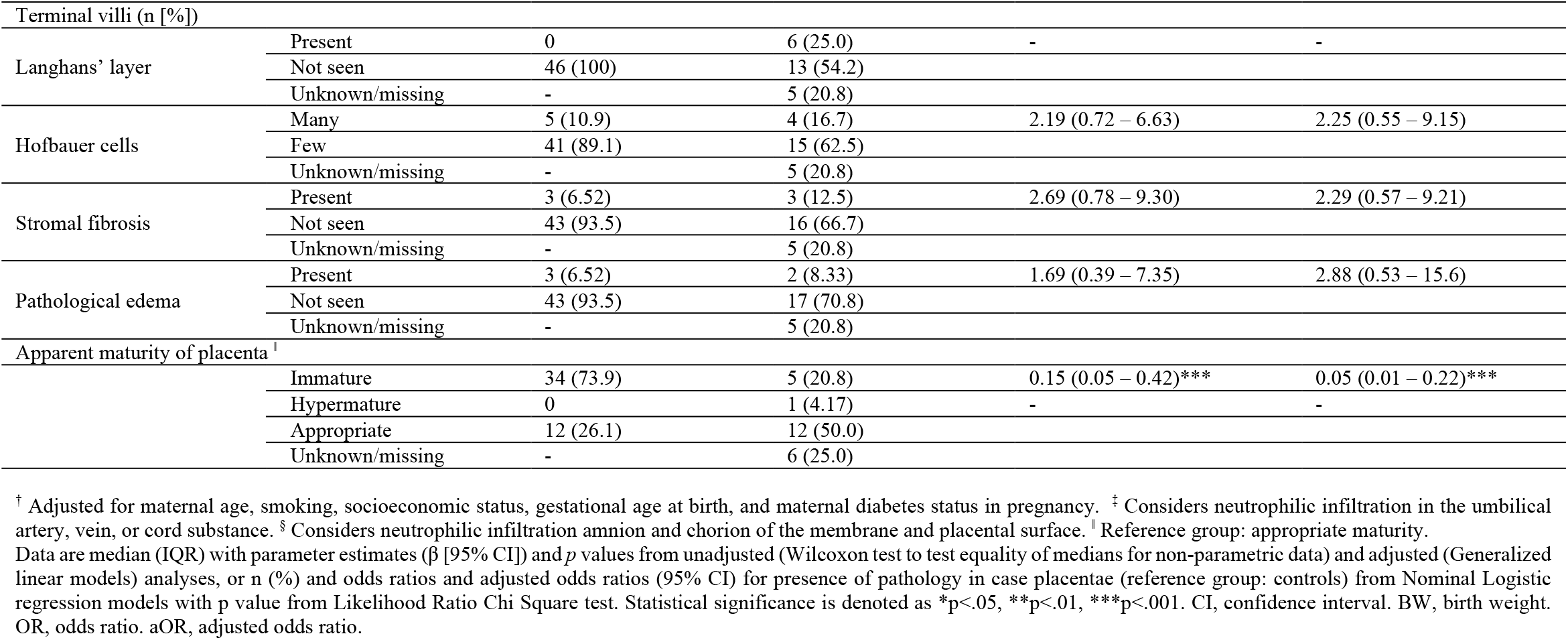
Associations between isolated neural tube defects and infant and placental characteristics and pathologies in cases and controls born preterm (<37 weeks’ gestation).

**Supplementary Table 5.** Associations between isolated neural tube defects and placental characteristics and pathologies in infants born term (≥37 weeks’ gestation).

| | | Control (n=102) | Case (n=50) | Unadjusted $\beta$ (95% CI) | Adjusted $\beta$ (95% CI) <sup>†</sup> |
| --- | --- | --- | --- | --- | --- |
| Infant characteristics |  |  |  |  |  |
| zWeight |  | -0.4 (-0.9 – 0.6) | -0.4 (-0.9 – 1.2) | 0.02 (-0.2 – 0.3) | 0.1 (-0.2 – 0.3) |
| Placental anthropometry |  |  |  |  |  |
| Weight (g) |  | 450 (385 – 504) | 391 (331 – 457) | -18.8 (-36.6 – -1.04)** | -8.78 (-28.0 – 10.4) |
| zWeight |  | -1.74 (-3.45 – -0.66) | -2.55 (-3.80 – -1.56) | -0.29 (-0.77 – 0.18) | 0.07 (-0.17 – 0.32) |
| Largest diameter (cm) |  | 19.5 (18.0 – 20.0) | 18.0 (17.0 – 20.0) | -0.51 (-0.86 – -0.16)** | -0.39 (-0.77 – 0.001) |
| Smallest diameter (cm) |  | 17.0 (16.0 – 18.0) | 16.0 (14.0 – 17.0) | -0.51 (-0.84 – -0.18)** | -0.23 (-0.58 – 0.12) |
| Thickness (cm) |  | 22.0 (20.0 – 25.0) | 23.0 (20.0 – 25.0) | 0.41 (-0.47 – 1.28) | 0.64 (-0.33 – 1.61) |
| Surface area (cm<sup>2</sup>) |  | 264 (226 – 290) | 214 (187 – 268) | -13.2 (-21.5 – -4.86)** | -7.47 (-16.5 – 1.55) |
| Birth weight (g):Placental anthropometry ratios |  |  |  |  |  |
| BW:Placental weight (g) |  | 7.27 (6.73 – 8.11) | 7.69 (6.71 – 8.86) | 0.29 (-0.02 – 0.61) | 0.21 (0.08 – 0.50) |
| BW:Largest placenta diameter (cm) |  | 173 (151 – 185) | 178 (160 – 197) | 5.70 (0.11 – 11.3) | 6.49 (1.13 – 11.8)* |
| BW:Smallest placenta diameter (cm) |  | 195 (179 – 214) | 201 (188 – 222) | 5.93 (-0.44 – 12.3) | 4.61 (-1.16 – 10.4) |
| BW:Placental surface area (cm<sup>2</sup>) |  | 12.6 (11.6 – 14.2) | 14.7 (12.3 – 16.3) | 0.90 (0.34 – 1.46)** | 0.71 (0.17 – 1.24)* |
| Placental pathology |  | Control (n=102) | Case (n=50) | Odds of pathology in term case placentae (vs. control)<br>OR (95% CI) | aOR (95% CI) <sup>†</sup> |
| Umbilical cord (n [%]) |  |  |  |  |  |
| Cord edema | Present | 2 (1.96) | 2 (4.00) | 2.15 (0.52 – 8.96) | 1.88 (0.22 – 15.9) |
|  | No description | 99 (97.1) | 46 (92.0) |  |  |
|  | Unknown/missing | 1 (0.98) | 2 (4.00) |  |  |
| Single umbilical artery | Yes | 0 | 5 (10.0) | - | - |
|  | No | 102 (100) | 43 (86.0) |  |  |
|  | Unknown/missing | - | 2 (4.00) |  |  |
| Neutrophilic infiltration <sup>‡</sup> | Any | 9 (8.82) | 5 (10.0) | 1.26 (0.50 – 3.20) | 1.58 (0.50 – 5.00) |
|  | Not seen | 93 (91.2) | 41 (82.0) |  |  |
|  | Unknown/missing | - | 4 (8.00) |  |  |
| Membranes & fetal surface (n [%]) |  |  |  |  |  |
| Cysts | Any | 5 (4.90) | 2 (4.00) | 0.86 (0.21 – 3.57) | 0.12 (0.003 – 6.02) |
|  | Not seen | 97 (95.1) | 45 (90.0) |  |  |
|  | Unknown/missing | - | 3 (6.00) |  |  |
| Neutrophilic infiltration <sup>§</sup> | Any | 20 (19.6) | 8 (16.0) | 0.86 (0.40 – 1.86) | 1.08 (0.42 – 2.75) |
|  | Not seen | 82 (80.4) | 38 (76.0) |  |  |
|  | Unknown/missing | - | 4 (8.00) |  |  |
| Maternal surface (n [%]) |  |  |  |  |  |
| Infarcts | Any | 21 (20.6) | 13 (26.0) | 1.43 (0.76 – 2.72) | 1.73 (0.80 – 3.72) |
|  | None | 81 (79.4) | 35 (70.0) |  |  |
|  | Unknown/missing | - | 2 (4.00) |  |  |
| Calcification | Any | 59 (57.8) | 24 (48.0) | 0.73 (0.41 – 1.29) | 0.73 (0.36 – 1.48) |
|  | None | 43 (42.2) | 24 (48.0) |  |  |
|  | Unknown/missing | - | 2 (4.00) |  |  |
| Syncytial knots | Excessive | 2 (1.96) | 1 (2.00) | 1.30 (0.18 – 9.56) | 4.30 (0.43 – 42.8) |
|  | Normal | 91 (89.2) | 35 (70.0) |  |  |
|  | Unknown/missing | 9 (8.82) | 14 (28.0) |  |  |
| Terminal villi (n [%]) |  |  |  |  |  |
| Langhans' layer | Present | 0 | 2 (4.00) | - | - |
|  | Not seen | 102 (100) | 44 (88.0) |  |  |
|  | Unknown/missing | - | 4 (8.00) |  |  |
| Hofbauer cells | Many | 2 (1.96) | 5 (10.0) | 6.10 (2.37 – 15.7)** | 7.78 (2.46 – 24.6)*** |
|  | Few | 100 (98.0) | 41 (82.0) |  |  |
|  | Unknown/missing | - | 4 (8.00) |  |  |
| Stromal fibrosis | Present | 7 (6.86) | 6 (12.0) | 2.04 (0.86 – 4.83) | 2.98 (1.18 – 7.53)* |
|  | Not seen | 95 (93.1) | 40 (80.0) |  |  |
|  | Unknown/missing | - | 4 (8.00) |  |  |
| Pathological edema | Present | 9 (8.82) | 9 (18.0) | 2.51 (1.21 – 5.23)* | 3.62 (1.48 – 8.82)** |
|  | Not seen | 93 (91.2) | 37 (74.0) |  |  |
|  | Unknown/missing | - | 4 (8.00) |  |  |
| Apparent maturity of placenta<sup>†</sup> |  |  |  |  |  |
|  | Immature | 0 | 0 | - | - |
|  | Hypermaturation | 5 (4.90) | 8 (16.0) | 4.15 (1.92 – 8.99)** | 5.73 (2.40 – 13.7)*** |
|  | Appropriate | 96 (94.1) | 37 (74.0) |  |  |
|  | Unknown/missing | 1 (0.98) | 5 (10.) |  |  |

<sup>†</sup> Adjusted for maternal age, smoking, socioeconomic status, gestational age at birth, and maternal diabetes status in pregnancy. <sup>‡</sup> Considers neutrophilic infiltration in the umbilical artery, vein, or cord substance. <sup>§</sup> Considers neutrophilic infiltration amnion and chorion of the membrane and placental surface. <sup>†</sup> Reference group: appropriate maturity. Continuous data are median (IQR) with parameter estimates ( $\beta$ [95% CI]) and *p* values from unadjusted (Wilcoxon test to test equality of medians for non-parametric data) and adjusted (Generalized linear models) analyses, or *n* (%) and odds ratios and adjusted odds ratios (95% CI) for presence of pathology in case placentae (reference group: controls) from Nominal Logistic regression models with *p* value from Likelihood Ratio Chi Square test. Statistical significance is denoted as \**p*<.05, \*\**p*<.01, \*\*\**p*<.001. CI, confidence interval. BW, birth weight. OR, odds ratio. aOR, adjusted odds ratio

**Supplementary Table 6.**
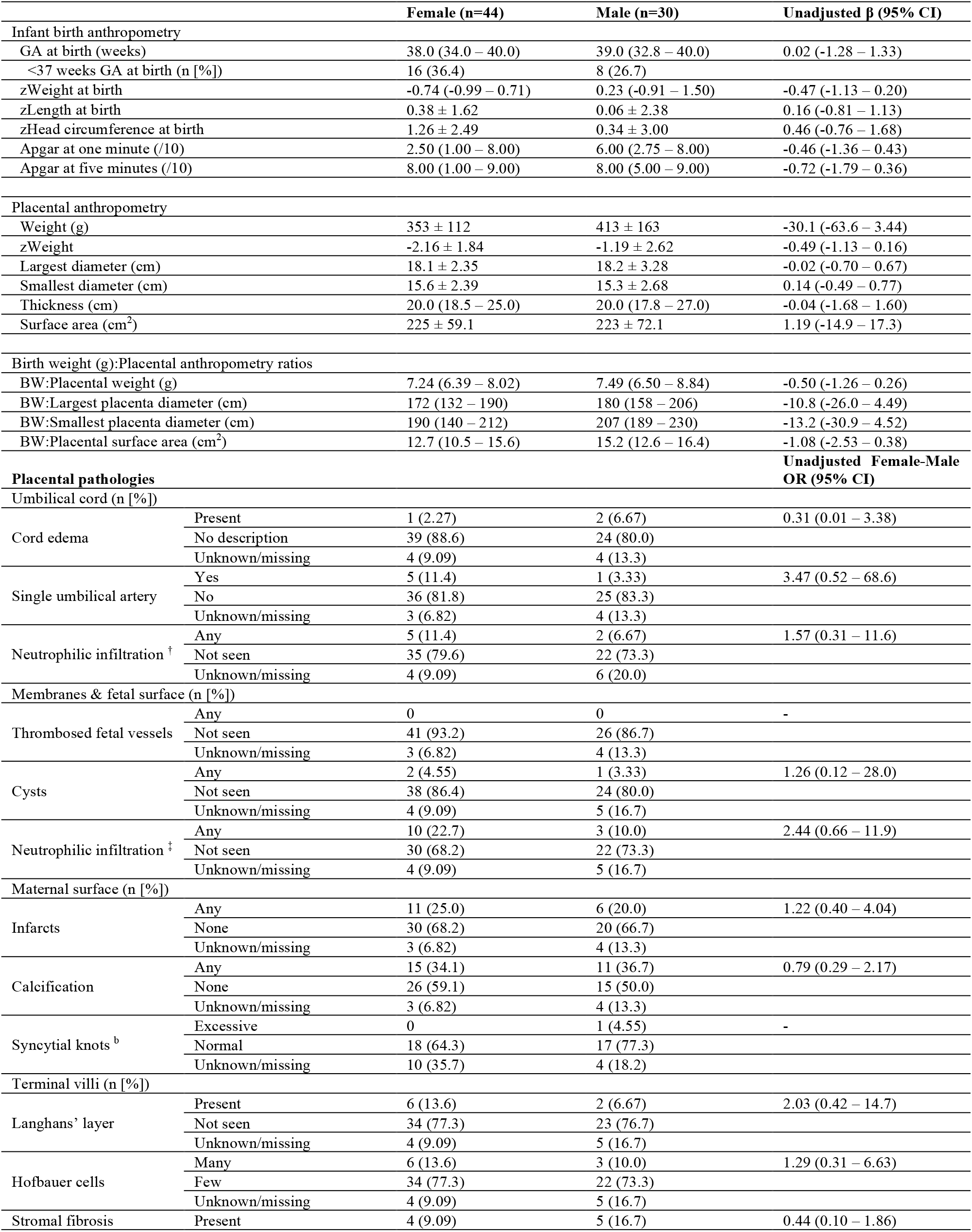
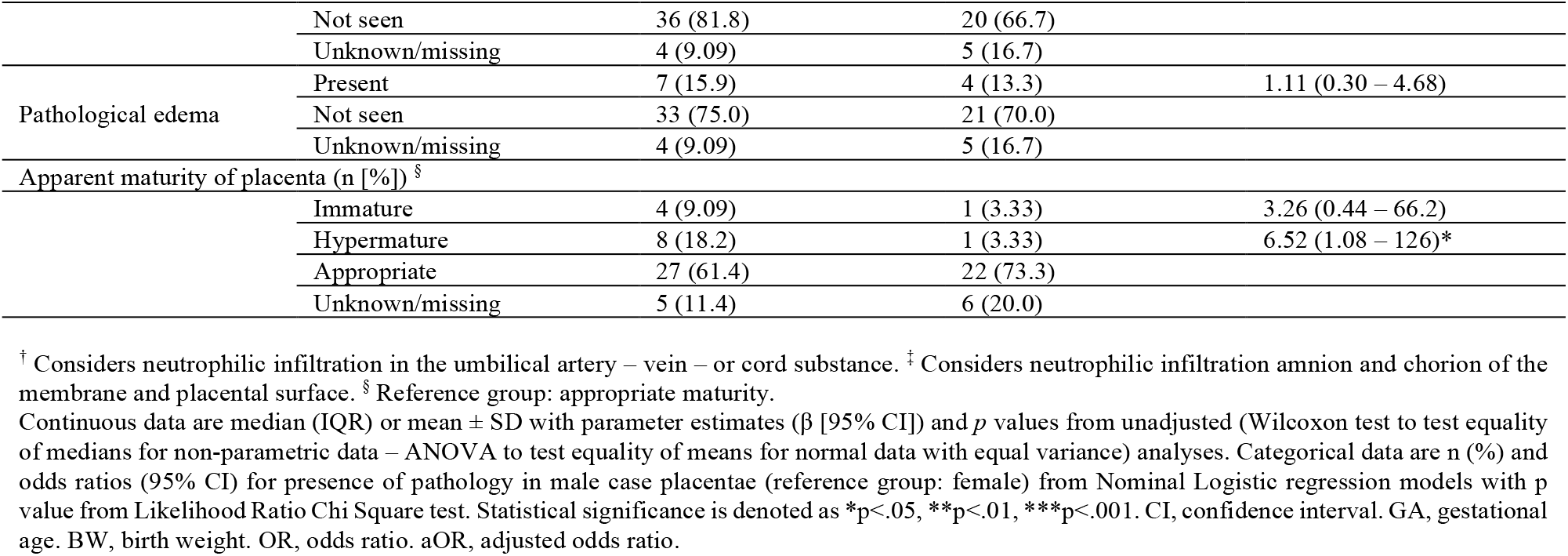
Infant birth characteristics and placental anthropometry and pathologies in female vs. male infants with an isolated NTD, Collaborative Perinatal Project (1959-1974).

## Notes

### Competing Interest Statement

The authors have declared no competing interest.

### Funding Statement

No external funding was received to support this study.

### Author Declarations

Carleton University Research Ethics Board (REB) exempts publicly available and anonymous data from REB review.

